# Dentine markers of pre/early postnatal lead exposure links with brain, cognitive, and behavioral outcomes in adolescents

**DOI:** 10.64898/2026.05.26.26354134

**Authors:** Andrew T Marshall, Eric Kan, Shana Adise, Maximilian König, Rob McConnell, Mauro Martinez, Vishal Midya, Manish Arora, Elizabeth R Sowell

## Abstract

Lead is a toxic metal ubiquitous in our environment. While dramatic reductions in lead sources have paralleled equivalent decreases in lead-poisoning rates, chronic lead exposure remains a critical public health concern. Childhood lead exposure (at its lowest levels) is linked to changes in cognitive development but less is known about lead’s effects on children’s brain structure, especially as a result of in utero exposure. We measured prenatal and early-postnatal lead exposure in shed deciduous teeth of 448 9- and 10-year-old children (from 20 United States cities) and linked those lead levels to childhood brain structure, cognition/behavior, and neighborhood- and family-level socioeconomic characteristics. Here we show negative associations between tooth-lead levels and the thickness of the brain’s cortex, particularly in regions linked to language processing. With increasing tooth-lead levels, children of lower-income (versus higher-income) families showed steeper declines in receptive vocabulary. Caregiver-reported behavioral problems exhibited similar associations. With in utero exposure linked to adverse neurodevelopmental outcomes (well before lead exposure and its risks are evaluated by healthcare professionals), prenatal screening of maternal lead levels/exposure, coupled with recommended strategies to reduce its placental transmission, may help reduce lead’s effects on future generations.

## Introduction

For two million years, hominids have been exposed to lead (Pb),^1^ an element recognized as a neurotoxicant (i.e., “one that makes the mind give way”) approximately 2,000 years ago by ancient Greek physician Dioscorides.^2^ Yet, society has only recently intervened to reduce lead’s neurotoxicity and its adverse effects on cognition, behavior,^3^ neuroanatomy,^4^ cardiovascular and respiratory outcomes,^5,6^ and pathogeneses of neuropsychiatric and neurodegenerative disorders.^7–9^ By limiting maximum lead content of housepaint, removing lead from on-road-vehicle gasoline, and building lead-free infrastructure for water-supply networks,^10^ lead-poisoning rates in the US have declined.^11^ But its effects persist, notably for the estimated 50% of the current US population exposed to high lead levels in early childhood.^12^

Removing direct lead sources from the environment does not necessarily excise the lead deposits^13^ from inside/outside the household^14^ or the body^15^ due to intergenerational transmission of lead exposure and its risks.^16^ Fetuses are subjected to (1) acute lead exposure (from the pregnant person during pregnancy) and (2) the remobilization of lead (stored in bone) into the pregnant person’s bloodstream,^15,17^ the latter of which increases during and after pregnancy.^18–20^ Typically, maternal and/or umbilical-cord blood are collected to assess prenatal exposure despite being indirect, incomplete measures of exposure through gestation.^21^ Instead, deciduous (“baby”, primary) teeth, long used to quantify individuals’ *total* lead burden,^22^ have (through technological advancements) emerged as effective, validated biospecimens for directly analyzing prenatal versus postnatal lead exposure on a temporally granular level.^23,24^ Deciduous teeth grow in incremental patterns, forming layers that can date prenatal/postnatal deposition similarly to the way tree rings identify environmental conditions.^25^ Tooth-lead levels (1) are negatively associated with cognitive, behavioral, psychiatric, and academic outcomes^26–35^ and (2) reflect past exposure during specific developmental windows,^23,24^ which may have greater effects on the central nervous system than current/recent exposure does (as conveyed by blood-lead levels).^29,36^ Well-established trajectories of tooth development, beginning in the second trimester in utero, coupled with technological advancements in tooth analysis,^23,24^ permit partitioning prenatal versus postnatal tooth-lead levels into by-week/by-trimester exposure levels^23,24^ and how they relate to adolescent/adult outcomes.^33–35,37^ However, there are no data on how prenatal and early-postnatal tooth-lead levels are associated with brain structure in today’s children.

Attributing any outcome to greater lead exposure is complicated by comparable effects that low socioeconomic status (SES) has,^38^ especially as (1) those at greater risks of exposure tend to have lower SES^39^ and (2) SES moderates Pb’s effects.^40,41^ Greater lead exposure in low-income children promotes further disadvantage, accentuating existing disparities.^42^ Parsing contributions of prenatal versus postnatal Pb exposure to brain and cognitive development, independent of SES, could aid in implementing simple interventions to reduce lead levels, which may lower rates of delinquent, antisocial, and criminal behavior,^43^ elevate academic performance,^43^ and increase lifetime-earnings estimates.^44^ However, most lead studies involve blood-lead measurements in small samples sizes, recruited from a limited geography at higher risk of lead exposure. In 9,712 9- and 10-year-old children in the Adolescent Brain Cognitive Development^SM^ Study cohort (hereafter, “ABCD”), we showed that children of lower-income (versus higher-income) households showed more pronounced decreases in brain structure and cognition with increases in lead-exposure risk,^45^ but, until now, we had no biomarkers of lead exposure in any of ABCD’s participants. To our knowledge, there are no previously published studies examining impacts of prenatal and postnatal lead exposure on brain structure *and* cognition in typically developing children across wide socioeconomic and geographic spectra. We hypothesized that prenatal and postnatal tooth-lead levels would be negatively associated with estimated, validated, geocoded, sociodemographic risk, neuroanatomical structure, cognitive performance, and caregiver-reported behavioral problems and that these associations would be more pronounced in children from low-income families.

## Results

### The Adolescent Brain Cognitive Development^SM^ Study (ABCD)

ABCD is the largest long-term study of brain development and child health in the United States. Between 2016-2018, it enrolled 11,878 9- and 10-year-old youth at 21 study sites (in 20 cities) for its 10-year duration.^46^ ABCD data are regularly made public via National Institutes of Health dissemination mechanisms. A total of 15,801 deciduous teeth from 4,708 youth participants were donated, with an initial goal to analyze how prenatal and postnatal heavy-metal and neurotoxicant exposures were associated with developmental outcomes in childhood/adolescence. Here, we present tooth-lead levels of 448 ABCD participants, randomly sampled from the larger repository with respect to household income, lead-exposure risk, and ABCD study site; baseline-appointment neuroimaging, cognitive, and behavioral data are available for 437 (97.5%), 440 (98.2%), and 448 (100%) of those participants, respectively. Tooth-lead levels in the deciduous-tooth dentine were collected using laser ablation inductively coupled plasma mass spectrometry (LA-ICP-MS).^23,24^ Tooth-lead exposure was operationally defined as the value (counts) of isotope ^208^Pb, background subtracted, normalized to the internal standard calcium-43 to control for variations in mineral content (i.e., ^208^Pb:^43^Ca ratios), and corrected to external standards (NIST610).

50.4% of our sample was assigned female sex-at-birth. By design, we targeted similarly sized groups across both (1) household-income brackets [Low (<$50K/year): *n*=147; Medium ($50-$100K/year): *n*=145; High (>$100K/year): *n*=156] and (2) estimated risks of lead exposure, per a census-tract-level map by the Washington State Department of Health (Table S1),^47^ the latter of which is a weighted composite of both poverty rates and age of housing by census tract. At ABCD’s baseline appointment, our sample was ∼9.9 years old (SD=0.6; range=9.0-11.0); youth’s caregivers tended to be highly educated, much like the overall ABCD population,^48^ with 69.4% (*n*=311) having completed at least a bachelor’s degree.

### Profiles of Prenatal and Postnatal Lead Exposure

ABCD was designed to collect youth data longitudinally from 9-10 to 19-20 years old, so participants’ socioeconomic data around birth are unavailable. However, because we planned to evaluate how prenatal/postnatal tooth-lead levels were associated with developmental outcomes in early adolescence (i.e., ABCD’s baseline), our planned analyses would account for youth demographics at this same baseline appointment. To better understand our sample’s tooth-lead levels, we evaluated if (and how) prenatal and early-postnatal tooth-lead levels varied by participants’ sociodemographics in late childhood/early adolescence, controlling for sex and ABCD site. All tooth types were analyzed (central incisors: *n*=48; lateral incisors: *n*=67; canines: *n*=123; 1^st^ molars: *n*=126; 2^nd^ molars: *n*=84), with data ranging from –21 to +55 weeks-since-birth (WSB), resulting in 22,325 ^208^Pb:^43^Ca datapoints log-transformed for analysis; 57 datapoints (0.26%) were removed for being ≤ 0. Given differences in dentition development by tooth type,^49^ there were differences in availability of participants’ data by WSB^50^ (Figures S1-S2); thus, we have primarily reported on tooth-lead levels from condensed WSB windows. [Analytically, when tooth lead was the modeled outcome, in which repeated tooth-lead measures were accounted for using by-participant random intercepts, we used the more traditional alpha (□) criterion at 0.05; for other analyses, in which repeated, weekly measures of tooth lead served as a predictor for a single, long-format-repeated neuroanatomical, cognitive, or behavioral measure, in which we controlled for by-participant time-varying tooth-lead levels, we set the criterion (and false-discovery-rate [FDR] correction criterion) at a very strict 0.005^51^ to minimize Type-1 errors.]

Figure 1 shows tooth-lead levels from -20 to +43 WSBs, a range corresponding to when ≥25% of participants had tooth-lead data; the shaded box (-12 to +13 WSB), when ≥90% of participants had data. Tooth-lead levels decreased with annual household income (WSB_-12:+13_: partial correlation coefficient [*r_p_*]=-0.02, *p*=.036; WSB_-20:+43_: *r_p_*=-0.01, *p*=.044; Figure 1A), decreased with caregiver education (WSB_-12:+13_: *r_p_*=-0.02, *p*=.019; WSB_-20:+43_: *r_p_*=-0.02, *p*=.010; Figure 1B), and increased with greater geocoded risk of lead exposure (WSB_-12:+13_: *r_p_*s≥0.02, *p*s≤.008; WSB_-20:+43_: *r_p_*s≥0.02, *p*s≤.003; Figure 1C) (Tables S2-S5). Across ABCD’s study sites, there was no clear geographical pattern as to those who were at high lead-exposure risk (Figure 2A), but individuals’ mean tooth-lead levels were significantly correlated with census-tract-level Pb-exposure risk (Pearson’s *r*=.17, *p*<.001; Figure 2B), especially census-tract-level poverty rates (Pearson’s *r*=.18, *p*<.001; Figure 2D) versus age of housing (Pearson’s *r*=.12, *p*=.014; Figure 2C). Cross-sectionally, youth living in households with (1) lower incomes *and* (2) caregivers with less education in (3) high lead-risk census tracts (risk = 8-10) (*n*=34) had significantly greater tooth-Pb levels than those living in households with (1) higher incomes *and* (2) caregivers with more education in (3) low lead-risk census tracts (risk = 1-3) (*n*=58), *r_p_*s≥0.07, *p*s<.001 (Figure 1D) (Tables S6-S7).

**Figure 1.**
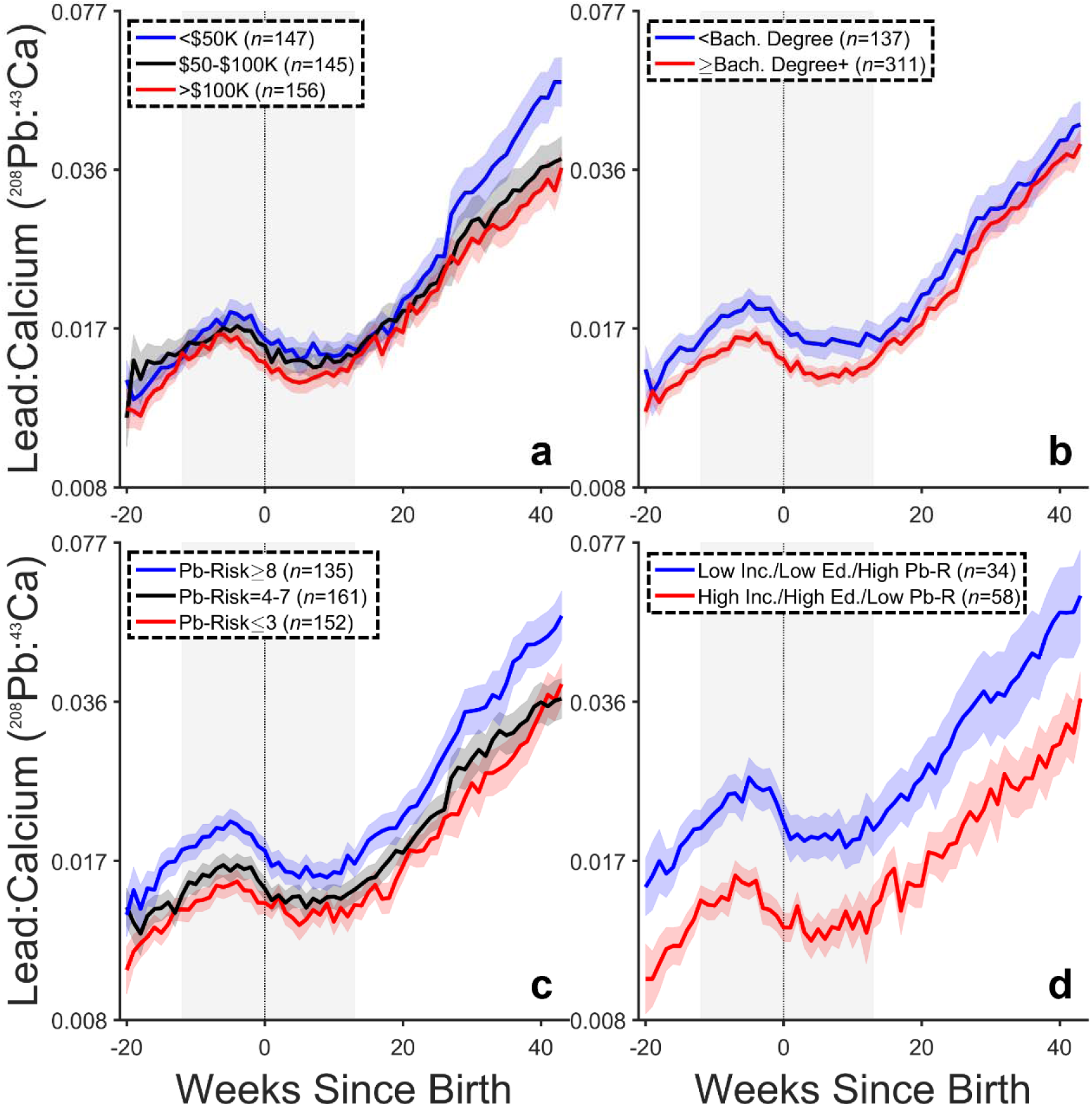
Prenatal and postnatal profiles of lead (Pb) exposure from tooth-Pb levels. Pb exposure was expressed relative to calcium (Ca) content in each tooth. The data shown represent mean lead exposure ± 1 standard error of the mean (SEM) as a function of week since birth (WSB), with negative WSBs representing the prenatal period and positive WSBs representing the postnatal period. Separate functions within each panel refer to participant sociodemographics when they were 9- to 10-years-old (**a**: Annual household income; **b**: Maximum caregiver education; **c**: Risk of Pb exposure; **d**: Sociodemographic intersections of **a**, **b**, and **c**). The size of each subgroup is shown. The shaded, gray box in each panel represents the -12 to +13 WSB window, in which at least 90% of participants had tooth data. High Inc. = annual household income was greater than $100,000/year (see **a**). Low Inc. = annual household income was less than $50,000/year (see **a**). Low Ed. = maximum caregiver education was less than a bachelor’s degree (see **b**). High Ed. = maximum caregiver education was at least a bachelor’s degree (see **b**). High Pb-R = risk of lead exposure was greater than or equal to 8 (see **c**). Low Pb-R = risk of lead exposure was less than or equal to 3 (see **c**).

**Figure 2.**
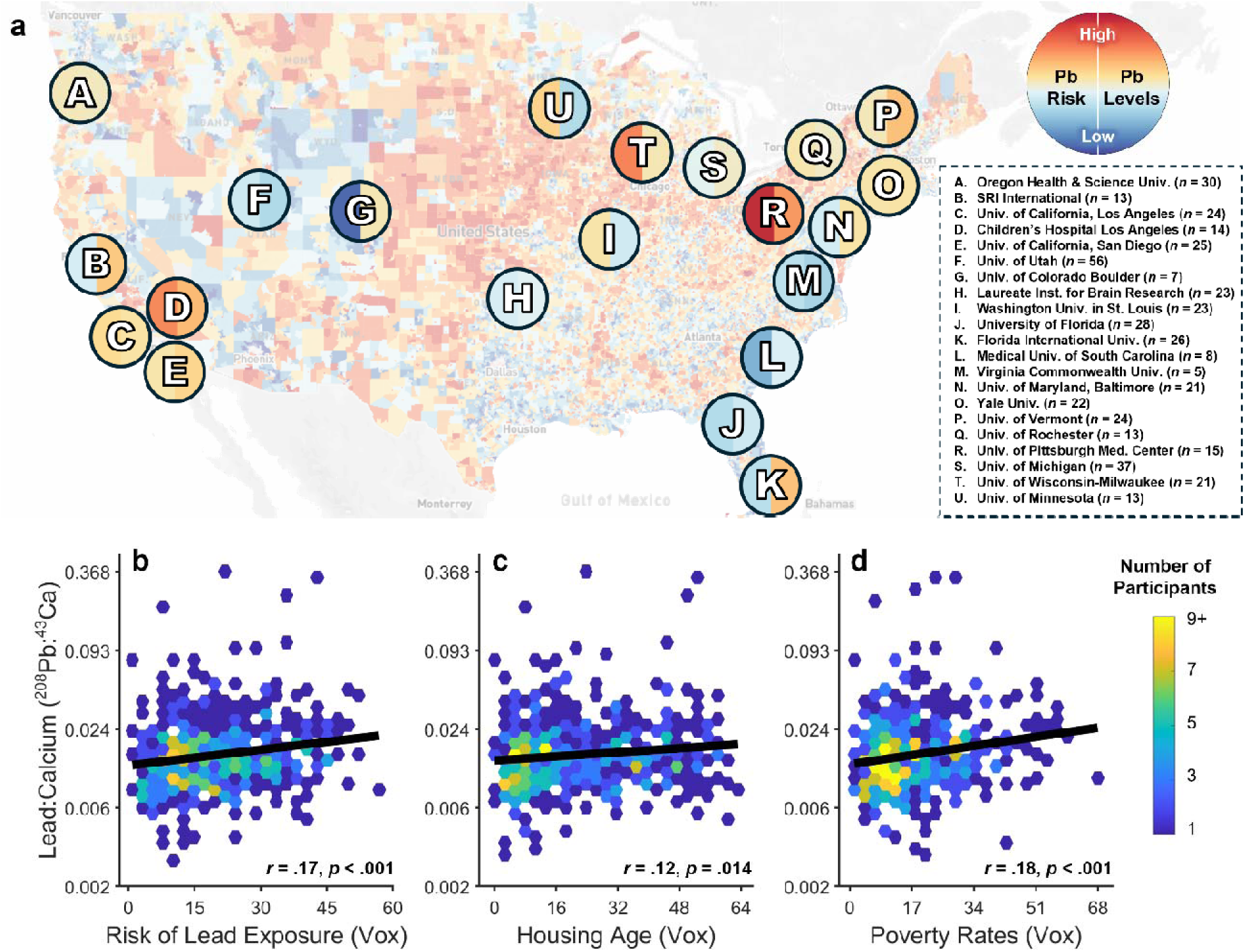
Relationship between risk of lead (Pb) exposure and prenatal and postnatal tooth lead levels. **(a)** The census-tract-level United States map is color-coded based on estimated risk of lead exposure given age of homes and poverty rates in that census tract, as estimated by a Washington State Department of Health metric published by Vox.^77^ Each lettered, bifurcated circle is approximately located at each of the 21 study sites in the Adolescent Brain Cognitive Development (ABCD) Study. The left half of each circle corresponds to the average risk of lead exposure across participants (*n*) from that study site in our sample; the right half, average tooth-lead levels. Lead exposure was expressed relative to calcium (Ca) content in each tooth. (**b**-**d**) Relationships between individuals’ mean tooth-lead levels and (**b**) risk of lead exposure and (c, d) its two components: (**c**) housing age and (**d**) poverty rates. Tooth-lead data were log-transformed. Data are shown as a hexbin-scatter plot, in which each hexagon is color-coded with respect to the number of individuals within that bin. The thick line is a simple regression line, with the strength of that relationship shown in each panel. Inst. = institute. Med. = medical. Univ. = university.

Past research has suggested that hand-to-mouth behaviors represent direct pathways to postnatal Pb ingestion.^52–54^ Per the American Academy of Pediatrics, this behavior will have started by 3-4 months postnatally.^55^ Beginning at +13 WSB (+3 months), there was a significant increase in tooth-lead levels, *r_p_*s≥0.50, *p*s<.001 (Tables S8-S11). Tooth-lead levels increased at significantly slower rates in youth living in homes with (1) higher household incomes, *r_p_*=-0.10, *p*<.001, and (2) caregivers with higher education levels, *r_p_*=-0.03, *p*=.004. Lead-exposure risk did not moderate these tooth-lead-level increases in this age range, *r_p_*=-0.004, *p*s≥.751.

### Cortical Structure, Cognition, and Behavior

ABCD’s extensive baseline battery (when participants were 9-10-years-old) included structural magnetic resonance imaging (MRI), cognitive tasks (i.e., NIH Toolbox), and caregivers’ reports on behavioral issues [i.e., Child Behavior Checklist (CBCL)]. We evaluated how overall (WSB_-12:+13_), prenatal (WSB_-12:0_), and postnatal tooth-lead levels (WSB_+1:+13_) related to regional cortical structure (thickness and surface, derived from acquired T_1_w MRI volumes using FreeSurfer v.7.1.1), cognition, and behavior, in which multiple individual tooth-lead levels were used to predict a neuroanatomical, cognitive, or behavioral outcome. These analyses controlled for participants’ sex-at-birth, age, household income, WSB, geocoded lead-exposure risk (given our past research^45^), ABCD study site/MRI scanner, and overall brain size (for cortical analyses) at baseline. Reductionist cognitive and behavioral analyses were performed to identify whether a particular cognitive subtask or class of caregiver-reported behavioral problems drove any significant associations between tooth-lead levels and broader composite cognitive or behavioral metrics.

#### Cortical structure

Prenatal and early-postnatal tooth-lead levels (WSB_-12:+13_; 11,220 tooth-lead-level observations across 437 participants) were more strongly, broadly, and consistently associated with early adolescent cortical thickness (vs. surface area) (Figure 3). Generally, greater tooth-Pb levels were linked to thinner cortices (Figures 3A–4, Table S16); the directionalities of corresponding surface-area relationships were more varied (Figure 3B, Table S36). The strongest cortical-thickness associations involved the bilateral middle-temporal cortex (left hemisphere [LH]: partial correlation coefficient [*r_p_*] = -0.09, *p*<.001; right hemisphere [RH]: *r_p_*=-0.11, *p*<.001), the bilateral supramarginal cortex (LH: *r_p_*=-0.09, *p*<.001; RH: *r_p_*=-0.09, *p*<.001), the bilateral inferior-parietal cortex (LH: *r_p_*=-0.08, *p*<.001; RH: *r_p_*=-0.08, *p*<.001), the LH pars orbitalis (*r_p_*=-0.09, *p*<.001), and the LH banks of the superior temporal sulcus (*r_p_*=-0.10, *p*<.001) (Figure 4). The inverse tooth-Pb – thickness relationships for the bilateral supramarginal (*r_p_*s≤-0.05, *p*s<.001), bilateral inferior-parietal (*r_p_*s≤-0.04, *p*s<.001), and bilateral middle-temporal cortices (*r_p_*s≤-0.08, *p*s<.001) were maintained when excluding multivariate outliers (Figure S3, Table S26).

**Figure 3.**
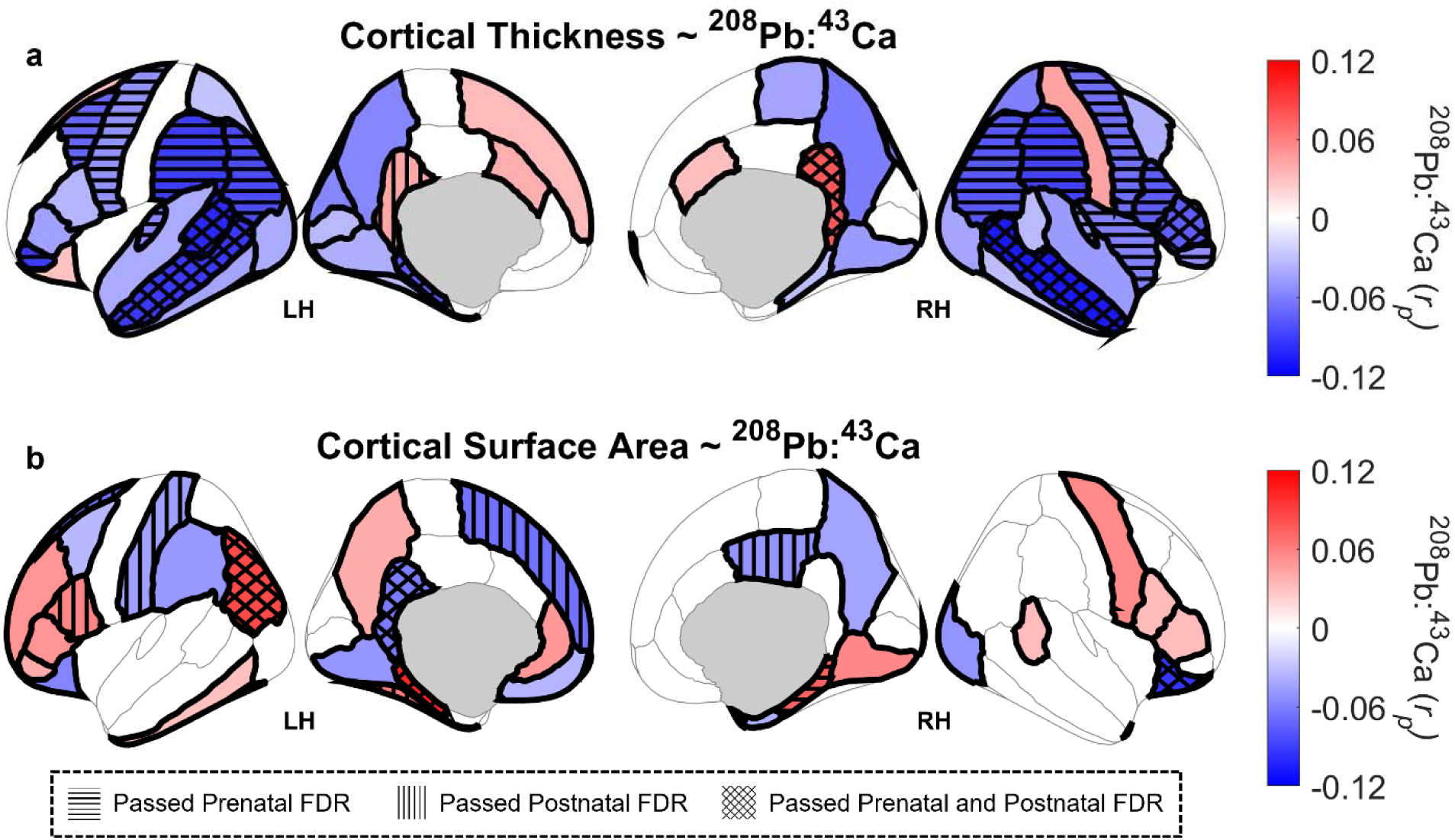
Associations between tooth-lead (Pb) levels and cortical structure. Bilateral cortical regions are color-coded based on the strength of the association between tooth-lead levels and cortical structure, controlling for sex, age, week-since-birth (WSB), household income, lead-exposure risk, Adolescent Brain Cognitive Development (ABCD) study site and magnetic resonance imaging scanner, and overall brain size (cortical thickness: whole-brain volume; cortical surface area: total surface area). Color-coding (per partial correlation coefficients, *r_p_*) is specific to analyses including weekly tooth-lead levels between -12 and +13 WSB, in which all tooth-lead levels within that 26-WSB range were included in analysis. Regions are shaded if those associations passed false-discovery-rate (FDR) correction; non-shaded regions indicate that the corresponding association did not pass FDR correction. Regions with a red (blue) shading indicate positive (negative) relationships between tooth-lead levels and cortical structure. Tooth-Pb levels were expressed relative to calcium (Ca) content in each tooth. Sensitivity analyses were performed for prenatal tooth-lead levels (-12:0 WSB) and postnatal tooth-Pb levels (+1:+13 WSB). Whether and how regions are shown as being hatched (i.e., horizontal, vertical, or crossed diagonal lines) indicate whether the tooth-lead – cortical structure relationships in pre- and/or postnatal analyses for that region also passed FDR correction. LH = left hemisphere. RH = right hemisphere.

**Figure 4.**
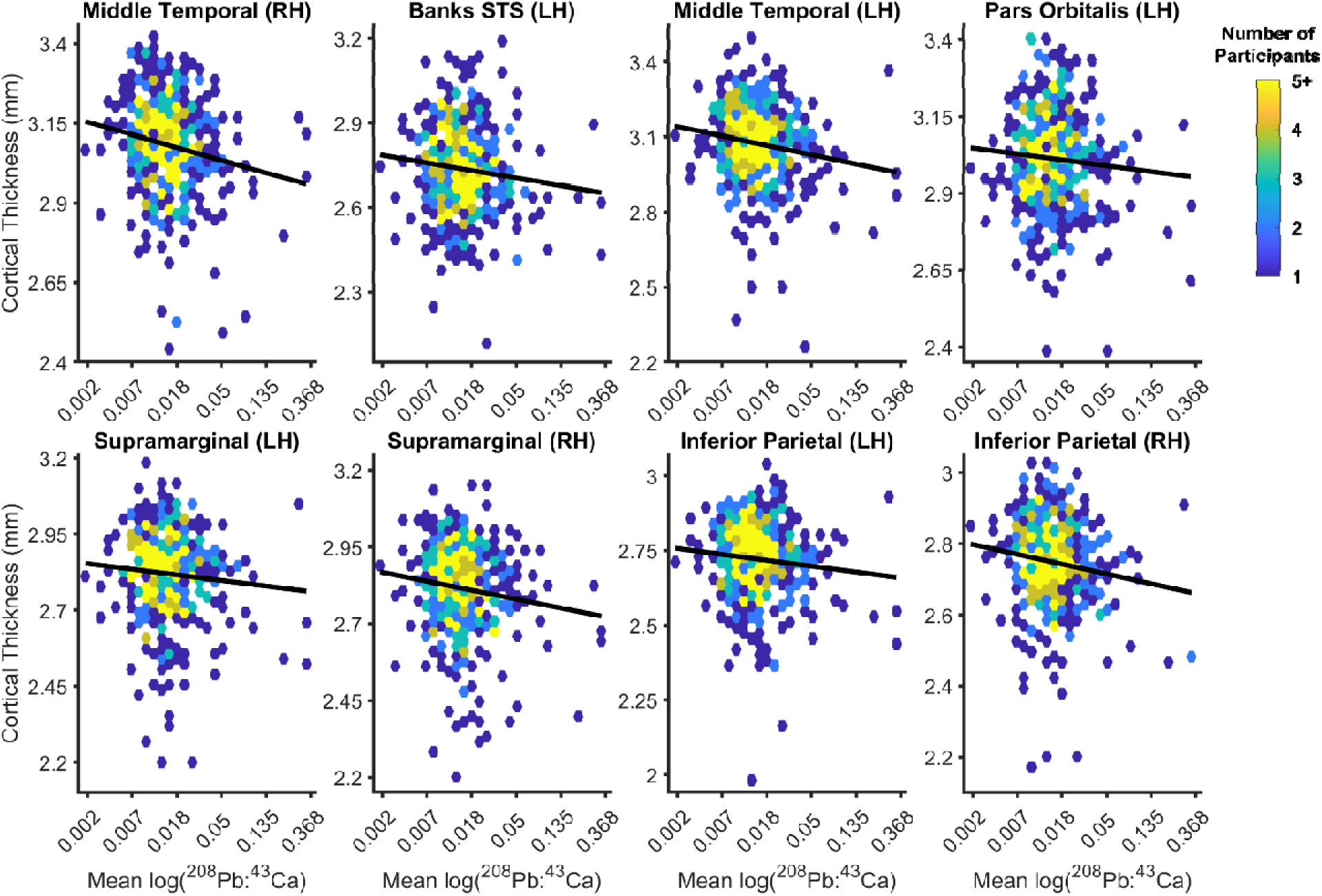
Strongest associations between prenatal and postnatal tooth-lead (Pb) levels and cortical thickness (in mm). The 8 regions shown correspond to 8 strongest associations in Figure 3a, with respect to the omnibus analyses. Lead exposure was expressed relative to calcium (Ca) content in each tooth; tooth-lead data were log-transformed, with these data referring to individuals’ mean tooth-Pb levels from -12 to +13 weeks since birth. Data are shown as a hexbin-scatter plot, in which each hexagon is color-coded with respect to the number of individuals within that bivariate bin. The thick line is a simple regression line for ease of interpretation. Banks STS = banks of the superior temporal sulcus. LH = left hemisphere. RH = right hemisphere.

Prenatal tooth-lead levels (WSB_-12:0;_ 5,567 tooth-lead-level observations across 437 participants) were significantly associated with cortical thickness of the bilateral middle-temporal, supramarginal, and inferior-parietal cortices (*r_p_*s≤-0.05, *p*s<.001); postnatal tooth-lead levels (WSB_+1:+13;_ 5,653 tooth-lead-level observations across 437 participants) were significantly associated with bilateral middle-temporal cortical thickness (*r_p_*s≤-0.05, *p*s<.001) (Figure 3A, Figures S4-S5, Tables S56 and S76), but the corresponding associations for the bilateral supramarginal and inferior-parietal cortices did not pass FDR correction (*r_p_*s≤-0.04, *p*s≤.002). As with main associations for cortical surface area, there was less consistency for how household income moderated associations between tooth-lead levels and both cortical metrics (Figures S6-S9; Tables S21, S31, S41, S51, S61, S71, S81, and S91).

#### Cognition and behavior

Greater tooth-lead levels were associated with lower NIH Toolbox composite scores (*n*=440), *r_p_*=-0.03, *p*<.001, which was primarily driven by reduced performance in crystallized cognition (*n*=444), *r_p_*=-0.05, *p*<.001, rather than fluid cognition (*n*=440), *r_p_*=-0.01, *p*=.116 (Tables S92-S94). The crystallized-cognition results were primarily driven by tooth-lead’s negative association with picture-vocabulary scores (receptive language), *r_p_*=-0.08, *p*<.001, rather than oral-reading-recognition scores (expressive language), *r_p_*=0.005, *p*=.600; greater household incomes were associated with weaker links between tooth-lead levels and picture vocabulary scores (Tooth-Lead×Household Income; Figure 5A), *r_p_*=0.07, *p*<.001 (Tables S95-S96). Both prenatal and postnatal tooth-lead levels were similarly associated with lower picture-vocabulary scores, *r_p_*s≤-0.05, *p*s≤.001, associations weakened at greater household incomes, *r_p_*s≥0.04, *p*s≤.001 (Tables S100-S101; Figure S10). These results were maintained when excluding multivariate outliers (tooth-lead association: *r_p_*=-0.10, *p*<.001; Tooth-Lead×Household Income interaction: *r_p_*=0.06, *p*<.001) (Table S102; Figure S10).

**Figure 5.**
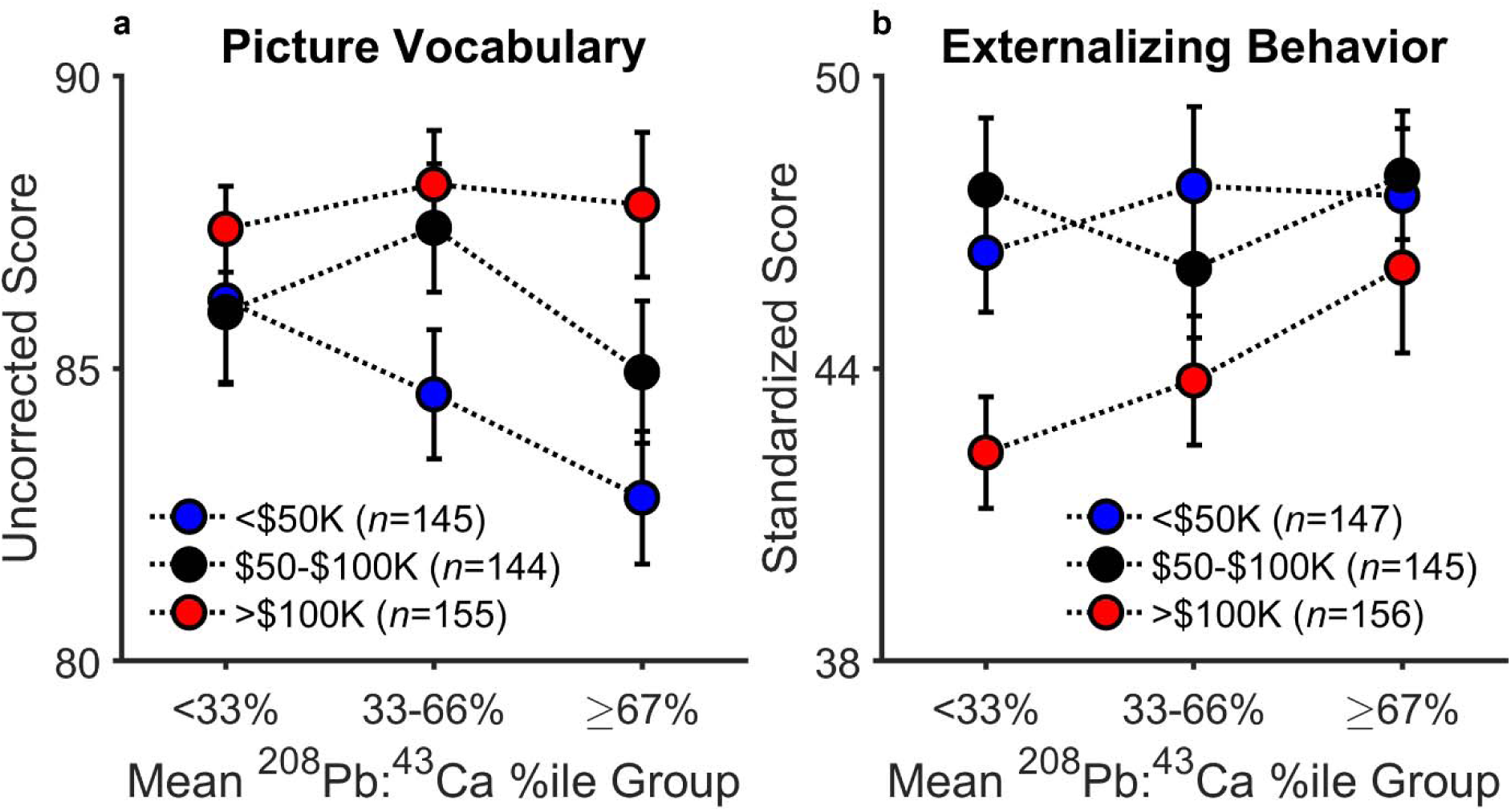
Household income moderated associations between tooth-lead (Pb) levels, cognition, and externalizing behavior. Lead exposure was expressed relative to calcium (Ca) content in each tooth; for ease of interpretation, participants’ mean tooth-lead levels (from -12 to +13 weeks since birth) were collapsed into 3 groups (i.e., the bottom third, middle third, and top third of mean tooth-lead levels). Within each panel, data are also split based on caregiver-reported annual household income, with the total numbers of participants in each subgroup shown. The corresponding analyses included individual tooth-lead levels by WSB and also controlled for sex, age, lead-exposure risk, and Adolescent Brain Cognitive Development (ABCD) study site. Uncorrected scores were analyzed for the NIH Toolbox (i.e., Picture Vocabulary) in line with our past research,^45^ while standardized scores were analyzed for the Child Behavior Checklist (i.e., Externalizing Behavior) to more closely approximate a normal distribution and for their clinical utility. Each datapoint reflects the mean ± 1 standard error of the mean (SEM).

Greater tooth-lead levels were also associated with more caregiver-reported total behavioral problems (*n*=448), *r_p_*=0.03, *p*=.002 (Table S97). While tooth-lead was positively associated with both externalizing and internalizing behavior, neither association was statistically significant after correction (externalizing: *r_p_*=0.03, *p*=.006; internalizing: *r_p_*=0.02, *p*=.044; Tables S98-S99). In contrast, greater household income was linked to more pronounced, positive associations with externalizing behavior, *r_p_*=0.04, *p*<.001 (Figure 5B), but not internalizing behavior, *r_p_*=0.02, *p*=.039. There were no tooth-lead – externalizing-behavior associations (or interactions involving household income) for prenatal or postnatal tooth-lead levels, |*r_p_*|s≤0.03, *p*s≥.046, nor when excluding multivariate outliers, |*r_p_*|s≤0.03, *p*s≥.007 (Tables S103-S105; Figure S10).

## Discussion

Lead exposure represents a largely preventable public health risk, which has been addressed over the past several decades (e.g., reducing/removing lead from various products); unfortunately, its effects (as with other neurotoxicants) can persist well beyond the actual exposure event(s). For instance, placental transmission of lead (potentially stored for years in a pregnant person’s bones) could, effectively, initiate a delayed exposure window for the pregnant person and their fetus.^56^ Thus, with respect to our sample, it is likely that lead exposure from the 20^th^ century may beget some of its repercussions in the 21^st^ century. However, by the time such exposure would be typically evaluated and addressed clinically,^57^ lead may have already done damage.^17,34,35,58^ Indeed, much of what is known about lead’s effects on the brain derive from (1) adult populations, (2) youth populations at elevated risks of exposure, and (3) studies of current exposure (i.e., blood-lead levels).^4,59–61^ Here, we presented, to our knowledge, the first report of prenatal and early-postnatal lead exposure’s effects on adolescent brain structure and cognitive/behavioral outcomes within a large, geographically and socioeconomically diverse United States sample, coupled with how tooth-lead levels relate to geocoded exposure risks. In doing so, we have provided data that may inform guidelines/recommendations for addressing new lead exposures along with dormant echoes from past ones (i.e., maternal bone-lead deposition).^58^ Given real possibilities of continued lead exposure (from infancy to adulthood), the body’s storage and redistribution of ingested lead, and the likelihood that even the smallest effects accumulate over time,^62^ our results potentially underestimate the true costs of lead exposure at the individual and population levels.^63^

First, we presented socioeconomic differences in tooth-lead levels: Higher SES was associated with lower overall and smaller postnatal increases in tooth-lead levels (Figure 1). Research of populations in Belgium and Spain similarly showed socioeconomic associations with cord-blood-lead levels.^64,65^ Moreover, as shown previously (and here), tooth-lead levels peaked prior to birth, followed by postnatal increases.^25,33,37^ While we were limited by the discrepant timing between the tooth-lead levels and socioeconomic data (i.e., tooth-lead levels around birth vs. socioeconomic conditions in early adolescence), the presence of differences (by SES, as observed here) may reflect the rich interconnectedness of lead and SES.^39^ Beyond the well-established links between lower SES and higher lead risk/exposure,^39^ Reuben, Caspi et al.^66^ showed that greater blood-lead levels in 11-year-olds in New Zealand were not only associated with those individuals’ lower SES in adulthood 27 years later but also greater reductions in SES relative to that of their parents (i.e., downward social mobility). Now, consider (1) that lead absorption rates range from 40-50% in children,^67^ (2) that >90% of the body’s lead burden is found in bone (i.e., a marker of cumulative exposure^22^), and (3) that bone lead’s contribution to blood Pb increases during/after pregnancy.^18–20^ These phenomena, along with possible consequences that youth lead exposure has in adulthood,^66^ suggest that the stratifying, transgenerational cycles of low-SES➔high-lead➔low-SES^42^ may begin in the next generation even before that next generation is born. We cannot establish causality between SES and tooth-lead levels, but past research, along with our data, offers clues to addressing such causal pathways. For instance, if the low-SES➔high-lead relationship exists prenatally, partly due to stored lead in the pregnant person’s bones, can the combination of prenatal screening,^58^ safe (if possible) chelating agents during pregnancy,^68^ dietary changes (e.g., calcium supplementation^69^), and/or lifestyle changes (smoking cessation^70^), reduce both placental and intergenerational lead transmission?

Next, we described the geographical expanse of tooth-lead levels in ABCD, along with corresponding census-tract-level risk factors (Figure 2). Whereas the lead-risk scores pertained to an address where participants may not have lived at birth, research has shown between-neighborhood socioeconomic consistency across individuals’ lifetime addresses.^71,72^ Regardless, while various lead-exposure “hotspots” have become evident through research and advocacy,^73,74^ there are non-zero lead-exposure risks elsewhere. Lead-risk scores are significantly associated with biomarkers of lead exposure,^45,75,76^ as shown here, further supporting the census-tract-level lead-risk map^47,77^ as a valid, publicly accessible, informational resource (https://www.vox.com/a/lead-exposure-risk-map). However, the relatively shallow relationships between lead risk (and its components) and tooth-lead levels also indicate that lead-exposure risk cannot fully predict lead exposure; the abilities of future efforts to improve such prediction remain to be seen.^73^

Even though the lead-risk map’s two-pronged composite score weighted housing age more than poverty rate, poverty rate was more strongly associated with tooth-lead levels here. While this may reflect some improvement in housing quality in older homes (i.e., there are now weaker correlations between housing age and presence of lead paint), it could instead be explained by changes in relevant exposure pathways: Assuming lead-ingestion likelihood is lower in pregnant persons than in infants, housing age (as a proxy for lead-paint risk) may matter less than it would for predicting pediatric lead exposure. Indeed, stronger relationships between tooth-lead levels and poverty rates are congruent with the aforementioned low-SES➔high-lead➔low-SES cycle, in which individuals of lower SES may be more likely to live in neighborhoods of lower SES, thereby elevating risks of lead exposure, which, if ingested, may lead to further decreases in SES in future generations.^42^ While addressing neighborhood-level lead risks presents its own challenges (i.e., environmental Pb remediation efforts have had mixed results^78^), neighborhood-poverty➔tooth-lead relationships may warrant targeted prenatal-screening efforts for those of (and living in neighborhoods with) lower SES.

Finally, we reported robust relationships involving tooth-lead levels, cortical structure, and cognitive/behavioral outcomes (Figures 3–5). Specifically, greater tooth-lead levels were broadly and robustly associated with thinner cortices, particularly in posterior temporal-parietal brain regions; more precisely, the bilateral middle temporal cortex (i.e., regions linked to language processing^79^). We know of two reports on lead exposure and cortical thickness in youth participants: Kim, Kim et al.^80^ showed no association between concurrent blood-lead levels and frontal-lobe cortical thickness in 150 6- to 17-year-olds, and Fornasier-Bélanger, Muckle et al.^59^ reported significant (albeit non-FDR-corrected) inverse associations between cord-blood-lead levels and frontal regions in 72 16- to 21-year-olds (their regions-of-interest did not include the temporal lobe). Also, in 191 8- to 14-year-old adolescents in Mexico City, Rechtman, Reichenberg et al.^37^ reported inverse relationships between total brain volume and postnatal (not prenatal) mixtures of nonessential (e.g., lead) and essential (trace) minerals/nutrients. Here, while despite the approximately 10-year lead-exposure – neuroimaging gap (i.e., intermediate factors may have contributed to such inverse relationships), our data (from a much larger cohort than these studies) corroborate research on the effects of lead exposure (and lead-exposure risk). In ABCD, we showed that greater risks of lead exposure are associated with reduced cortical surface area and volume (but not thickness), particularly in individuals living in homes with lower household incomes,^45^ which, while incongruent with the present data, may instead provide clues to both sets of results.

Notably, Bethlehem, Seidlitz et al.^81^ compiled 123,984 MRI scans to evaluate lifetime changes in human brain structure. They reported that, prenatally, cortical thickness is changing more rapidly than other neuroanatomical metrics. Thus, any changes to the prenatal environment may be most likely to affect cortical thickness. (Interestingly, using ABCD data, we have also shown that prenatal tobacco exposure has its strongest relationships with changes in cortical thickness versus surface area.^82^) Such temporality may help explain the negative yet nonsignificant relationships between childhood lead exposure and adulthood cortical thickness reported previously.^83^ If prenatal cortical thickness can account for variations in adolescent cortical thickness, then any lead-induced alterations in prenatal cortical development may be apparent during adolescent cortical development (Figures 3–4). Indeed, our sensitivity analyses supported this interpretation, given broader relationships between prenatal versus postnatal tooth-lead levels and cortical thickness (Figures S4-S5). While there were less consistent moderations of these relationships by household income (Figures S6-S9), it is possible that, despite separation in prenatal tooth-lead levels by SES (Figure 1), greater (or lower) household incomes do not serve to substantially weaken (or strengthen) relationships between prenatal lead exposure and adolescent cortical thickness, even if household income has an impact on relationships reflecting manifestations of neuroanatomical differences.

It has been widely reported that SES moderates relationships involving environmental exposures, neuroanatomy, cognition, and/or behavior.^84–86^ Here, we showed that greater prenatal and early-postnatal tooth-lead levels were associated with lower scores on receptive-vocabulary tests at 9- to 10-years-old, a relationship most pronounced in those living in lower-income households (Figure 5A). In other words, greater household incomes may be protective against neurotoxicant-associated deficits in receptive-vocabulary performance. In contrast, at greater tooth-lead levels, any protectiveness of higher household income on youth’s externalizing behavior may be weakened (Figure 5B). Past research has linked lead exposure, language processing, and behavioral problems; for example, across 42 young adults, Yuan, Holland et al.^87^ showed reduced activation (during a language-generation task) in the left (not right) middle temporal gyrus in those who had higher blood-lead levels as children. Overall, our data suggest that prenatal and early-postnatal lead exposure may alter trajectories of cortical-thickness development, as observed in early adolescence, which may contribute to differences in early-adolescent language processing (and externalizing behavior).

As noted above, there are limitations (some more reflective of the broader ABCD Study) to guide future research. Primarily, despite direct measures of prenatal and early-postnatal lead exposure in baby teeth, there are very limited data on ABCD children’s prenatal and early-postnatal environments. While having those data would not permit establishing causality, the temporal gap between lead exposure (per tooth-lead levels) and ABCD’s onset reflects possibilities that intervening events moderated the relationships observed here, a research question better addressed by future longitudinal studies investigating other critical developmental windows. Further, whereas ABCD was designed to be demographically diverse,^46^ it is not nationally (nor globally) representative. Our sample was more balanced with respect to household income (Table S1), but ABCD’s study sites in populated, relatively urban areas near universities/hospitals may explain the high education levels achieved by ABCD caregivers. However, studies showing relationships between prenatal tooth-lead levels and cognitive/mental-health outcomes in older adults^34,35^ and between tooth-lead levels and cognition/behavior in children^26–32^ suggests that such results are present across broader socioeconomic and geographic scales, as uniquely shown here.

In conclusion, the current study has provided, to our knowledge, a novel contribution to the literature, showing how (1) direct measures of prenatal and early-postnatal lead exposure across participants from (2) wide geographic and (3) socioeconomic spectra relate to (4) brain, behavioral, and cognitive outcomes at critical developmental timepoints within (5) the largest, longitudinal study of adolescent development in the United States. Ultimately, lead’s bone-to-blood remobilization during gestation (and lactation) can put the pregnant person and fetus at risk for lead exposure that occurred years earlier, lengthening lead’s effects through intergenerational transmission. There are no safe levels of lead exposure, and this study, like Gardella^58^, supports reconsideration for lead screening and appropriate treatment before the 6-month timepoint recommended now by the American Academy of Pediatrics,^57^ even for individuals not currently at elevated risks for lead exposure. In doing so, efforts to remediate lead from the environment may be more effective when considering the environment inside the body along with the one surrounding it.

## Online Methods

### Adolescent Brain Cognitive Development^□^ Study Cohort

The Adolescent Brain Cognitive Development□ Study (hereafter, “ABCD”) enrolled 11,878 9- and 10-year-old youth at 21 study sites (Figure 1) primarily using school-based enrollment.^46,88^ Our data (collected between September 2016 and October 2018) came from May 2026’s ABCD 7.0 data release (doi:10.82525/8f3w-5260; https://www.nbdc-datahub.org/abcd-release-7-0), which included initial/baseline-appointment data for 11,860 youth participants. Centralized IRB approval was obtained from UC San Diego. Study sites obtained approval from their IRBs. Caregivers provided written informed consent and permission; youth provided written assent. Data collection/analyses complied with all ethical regulations. Our report was structured in accordance with STROBE guidelines.^89^

### Current Sample

Here, we have described relationships between pre- and postnatal lead-exposure levels [as measured in shed deciduous (“baby”) teeth] and neuroanatomical, neurocognitive, and behavioral outcomes in early adolescence in ABCD youth participants. A total of 15,801 deciduous teeth from 4,708 youth participants were donated, with an initial goal to analyze how prenatal and postnatal heavy-metal and neurotoxicant exposures were associated with developmental outcomes in childhood/adolescence.^90^ Of the 4,708 participants who donated teeth, we sampled 500 participants based on baseline family income, race/ethnicity, baseline risk of lead exposure, and study site. The data for 448 of those participants are reported here (Table S1); the biospecimens of 52 participants did not pass quality control for sample quality. Our research questions were centered around estimated risk of lead exposure and household income, in line with our previous research,^45^ such that our goal was to have equivalent numbers of participants at different income and lead-risk levels. In turn, because these same questions did not depend on race/ethnicity and study site (both of which were incremental in ABCD’s overall target sample^46^), we allowed our sample to be distributed across race/ethnicity and study site in accordance with the breakdown of those factors in ABCD.

**Table S1.** Sample demographics.

|  |  | Race/Ethnicity |  |  |  |  | Total (%) |
| --- | --- | --- | --- | --- | --- | --- | --- |
|  |  | Asian | Black | Hisp. | Other | White |  |
| Family SES/Income Bracket | Low | 3<br>3 | 14<br>12 | 19<br>17 | 4<br>5 | 37<br>33 | 147 (33) |
|  | Mid | 4<br>3 | 12<br>14 | 16<br>15 | 4<br>4 | 37<br>36 | 145 (32) |
|  | High | 5<br>4 | 14<br>11 | 18<br>19 | 5<br>5 | 37<br>38 | 156 (35) |
| Total (%) |  | 22<br>(5) | 77<br>(17) | 104<br>(23) | 27<br>(6) | 218<br>(49) | 448<br>(100) |

\*Note: Within each Income × Race/Ethnicity cell, the number in the unshaded section refers to those at "low" risk of lead exposure; the number in the shaded section refers to those at "high" risk of lead exposure. Hisp. = Hispanic

For sampling, caregiver-reported annual household income level at baseline, reported on an ordinal 1-10 scale, was categorized into low (<$50,000/year), medium ($50,000-$100,000/year), and high brackets (>$100,000/year), as done previously.^45,91^ Youth race/ethnicity was identified by caregivers at baseline appointments, during which ABCD’s race/ethnicity ongoing distribution target was 5.3% Asian, 16.6% Black, 23.2% Hispanic, 5.4% Other/Mixed Race, and 49.5% White/Caucasian.^46^ Summarily, we aimed for evenly sized income-bracket groups, each of which would have a race/ethnicity breakdown more closely aligned with ABCD’s said corresponding target distribution.

Lead risk (range = 1-10) was based on a high-resolution, United States census-tract-level nationwide map, generated by the Washington State Department of Health, published by Vox.^47,77^ Each tract’s lead-risk score was a weighted average of two tract-specific American-Community-Survey-derived values: the ages of homes (weight = 0.58) and poverty rates (0.42), both of which are well-established correlates of lead exposure.^39,92^ These lead-risk scores have also been shown to be associated with blood-lead levels in children.^45,73,75,76^ Census tracts were determined based on caregiver-reported youth’s primary home address at baseline. For sampling, we aimed for half of each of the Income × Race/Ethnicity groups in our sample to be at a low risk (scores = 1-5) and half at a high risk (scores = 6-10).

At the time of sample generation, participants’ teeth were located at individual ABCD study sites, having yet to be transferred to a central location. Thus, across 10,000 iterations of random-sample generation, our sample (in line with the above criteria) aimed to minimize the variance of study-site-by-study-site differences in contribution and labor (i.e., to package and ship the teeth for central analysis) relative to the total number of participants and number of teeth donated at each study site. Table S1 shows the cross-tabulated distribution of our sample for household income, race/ethnicity, and lead risk; 50.4% of our sample was female. Figure 1 shows the number of participants in our sample from each site, along with average lead risk and tooth-lead levels for the sample’s participants at that site.

### Procedures

#### Laser ablation inductively coupled plasma mass spectrometry (LA-ICP-MS)

Shed teeth with carious lesions, metal restoration, developmental defects, or significant loss of enamel or dentine are excluded. To identify temporal developmental zones, the neonatal line is visualized to detect prenatally versus postnatally formed regions; parts of dentine (and enamel) formed at different times are identified according to the visible daily growth rings, following our published methods.^93,94^ Tooth sections are analyzed for metals at 35-µm resolution using laser ablation-ICP-MS (Agilent, 8900 ICP-MS) with extended range detection and a New Wave Research NWR-193 (ESI, USA) laser ablation unit with a 193 nm ArF excimer laser.^23,24,93–98^ This resolution relates to approximately 50 time points/tooth representing weekly exposure from 14 weeks gestation through one year postnatal age. Helium was used as the carrier gas from the laser ablation cell, mixed with argon via a T-piece before being introduced into the ICP-MS. System tuning was performed daily using NIST SRM 612 (trace elements in glass) to optimize sensitivity (maximize ion counts), minimize oxide formation (^232^Th16O^+^/^232^Th^+^, < 0.3%), and monitor fractionation (^232^Th^+^/^238^U^+^, 100 ± 5%). Tooth-Pb exposure was (1) operationally defined as the value (counts) of isotope ^208^Pb and quantified by (2) normalizing to tooth calcium (Ca) levels (^208^Pb:^43^Ca) (because mineral density varies within teeth and between tooth type and individuals) and (3) ICP-MS ion counts for metals:Ca are quantified by external calibration using NIST SRM 610, 612, and 614 glasses.

#### Neuroimaging

ABCD’s neuroimaging acquisition parameters, data collection, and processing procedures have been thoroughly described previously.^99,100^ We analyzed the thickness (mm) and surface area (mm^2^) of 68 bilateral cortical regions [derived using FreeSurfer v.7.1.1 Desikan-Killiany atlas on acquired T_1_w structural magnetic resonance imaging (sMRI) volumes].^101,102^ FreeSurfer Automatic Segmentation was used to derive whole-brain volume,^99^ which was also included in analyses.

#### NIH Toolbox

Data collection procedures are described in detail elsewhere.^103,104^ Briefly, NIH Toolbox tests show good convergent validity compared with gold standards of cognitive testing^105^: (1) the Picture Vocabulary Test (a measure of receptive language/crystallized cognition; Version 2.0, Ages 3+), (2) the Oral Reading Recognition Test (expressive language/crystallized cognition; Version 2.0, Ages 3+), (3) the Flanker Inhibitory Control and Attention Test (attention, executive function/fluid cognition; Version 2.0, Ages 8–11), (4) the List Sorting Working Memory Test (working memory/fluid cognition; Version 2.0, Ages 7+), (5) the Dimensional Change Card Sort Test (executive function/fluid cognition; Version 2.0, Ages 8–11), (6) the Pattern Comparison Processing Speed Test (processing speed/fluid cognition; Version 2.0, Ages 7+), and (7) the Picture Sequence Memory Test (episodic memory/fluid cognition; Version 2.0, Form A, Ages 8+).

The NIH Toolbox was administered on an iPad (∼25–30 min to complete), with all task’s instructions being read by the examiner, except for the Pattern Comparison Processing Speed Test (i.e., presented by an audio recording). All tasks were administered with the iPad in an upright position (∼45° degree angle), with some tasks incorporating a home-base “button” in front of the iPad, on which the participants would place their index finger between trials (Flanker Inhibitory Control and Attention Test, Dimensional Change Card Sort Test).

#### Child Behavior Checklist (CBCL)

The CBCL is a 113-item caregiver-completed questionnaire on their youth’s behavioral problems from the past 6 months,^106^ first completed at the baseline appointment.^104,107^ For each item, response options are: 0 = Not True, 1 = Somewhat/Sometimes True, 2 = Very True/Often True; “Don’t Know” and “Decline to Answer” options were also provided. Summary scores were computed (and standardized) for internalizing-behavior, externalizing-behavior, and total problems.

#### Demographics

Annual total household income was a 10-level ordinal factor (with response options also for “Don’t know” and “Refuse to answer:): Less than $5,000; $5,000 through $11,999; $12,000 through $15,999; $16,000 through $24,999; $25,000 through $34,999; $35,000 through $49,999; $50,000 through $74,999; $75,000 through $99,999; $100,000 through $199,999; and, $200,000 and greater. Maximum caregiver education level was a 21-level ordinal factor (with an additional response option for “Refused to answer”): Never attended/Kindergarten only; 1st grade; 2nd grade; 3rd grade; 4th grade; 5th grade; 6th grade; 7th grade; 8th grade; 9th grade; 10th grade; 11th grade; 12th grade; High school graduate; GED or equivalent; Some college; Associate degree: Occupational; Associate degree: Academic Program; Bachelor’s degree (ex. BA); Master’s degree (ex. MA); Professional School degree (ex. MD); and, Doctoral degree (ex. PhD). For analysis, caregiver education was collapsed into 5 levels: ≤12^th^ grade, no diploma; high school graduate / GED; some college, no degree / associate’s degree; bachelor’s degree; and, advanced degree.

The lead-risk map (range: 1-10) that was used for sampling was also used for most analyses. For some graphical depiction (and for correlation analyses between lead-exposure risk and tooth-lead levels), we re-coded these deciles along a more granular 0-100 scale using its components of housing age and poverty rates from the 2010-2014 American Community Survey 5-year estimates. Here, housing age is an estimate of the proportion of homes in each census tract with lead-based paint hazards^39^ (sample range = 0.1–63.2). Poverty rates refer to the proportion of individuals in that census tract below -125% of the poverty level (sample range = 1.3-67.9). In accordance with the original lead-risk map, we computed a weighted sum of the housing-rate proportion (weight = .58) and the poverty-rate proportion (weight = .42) (i.e., Lead Risk = .58*Housing + .42*Poverty), with higher values reflecting greater lead risks.

### Data Analysis & Statistics

All statistical tests were performed in MATLAB R2022b (The MathWorks, Natick, MA, USA), primarily using MATLAB’s Statistics and Machine Learning Toolbox (Version 12.4). Given the nature of our tooth-lead (multiple observations) and adolescent-outcome data (one observation), along with the structure of the corresponding analyses, the alpha (□) level for statistical significance was set at 0.05 when tooth-lead levels were the criteria; it was set at 0.005 when tooth-lead levels were the predictor.

#### Lead exposure

All tooth-lead levels by week-since-birth (WSB) are reported as normalized to calcium (Ca) levels in the tooth, to control for overall mineral content.^23^ All tooth-lead levels were log-transformed for analysis, simply referred to as *Tooth Lead* (or *Tooth-Lead Levels*) below; individual tooth-lead levels less than or equal to zero were removed prior to transformation (*n* = 57; 0.26% of the data). Five types of teeth were included in our analyses: central incisors (*n* = 48), lateral incisors (*n* = 67), canines (*n* = 123), 1^st^ molars (*n* = 126), and 2^nd^ molars (*n* = 84). Collapsed across tooth types, data were available between -21 and +55 WSB, with a total of 22,325 datapoints.

Due to differences in timing of coronal primary dentine completion by tooth type,^49,50^ data are not available for all teeth across this 77-week range. Figure S1 shows the percentage of participants with tooth-lead data at each WSB (i.e., lightly shaded bars), along with the contribution of tooth type to the stair-step-like patterning in data availability. Thus, our primary analyses focused on tooth-lead levels for WSBs in which at least 90% of participants had data (i.e., -12 to +13 WSB); a general linear mixed-effects model indicated that there were no main effects of tooth type on tooth-lead levels between -12 and +13 WSB, controlling for individual-participant levels [i.e., *Tooth Lead_-_*_12*:+*13_ ∼ *Tooth Type* + (1|*Participant*)], *F*(4, 11497)=0.96, *p*=.425 (Figure S2). When specified, different temporal windows were used to address specific research questions.

**Figure S1.**
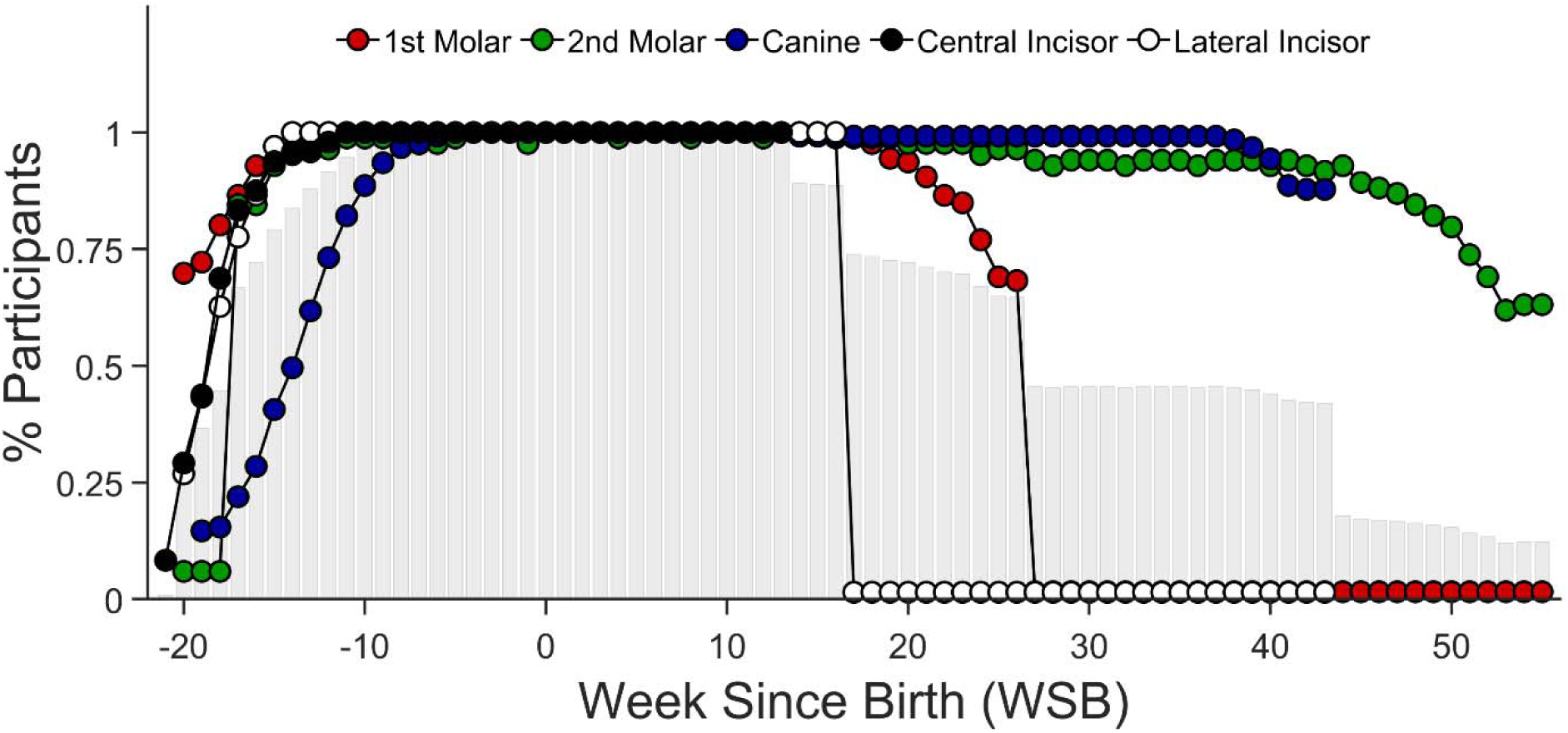
Tooth-data availability by week since birth (WSB). Lightly shaded, gray bars represent the percentage (%) of participants in the overall sample (*n* = 448) with data at each WSB. The datapoints show the percentage of participants by tooth type with tooth data at each WSB.

**Figure S2.**
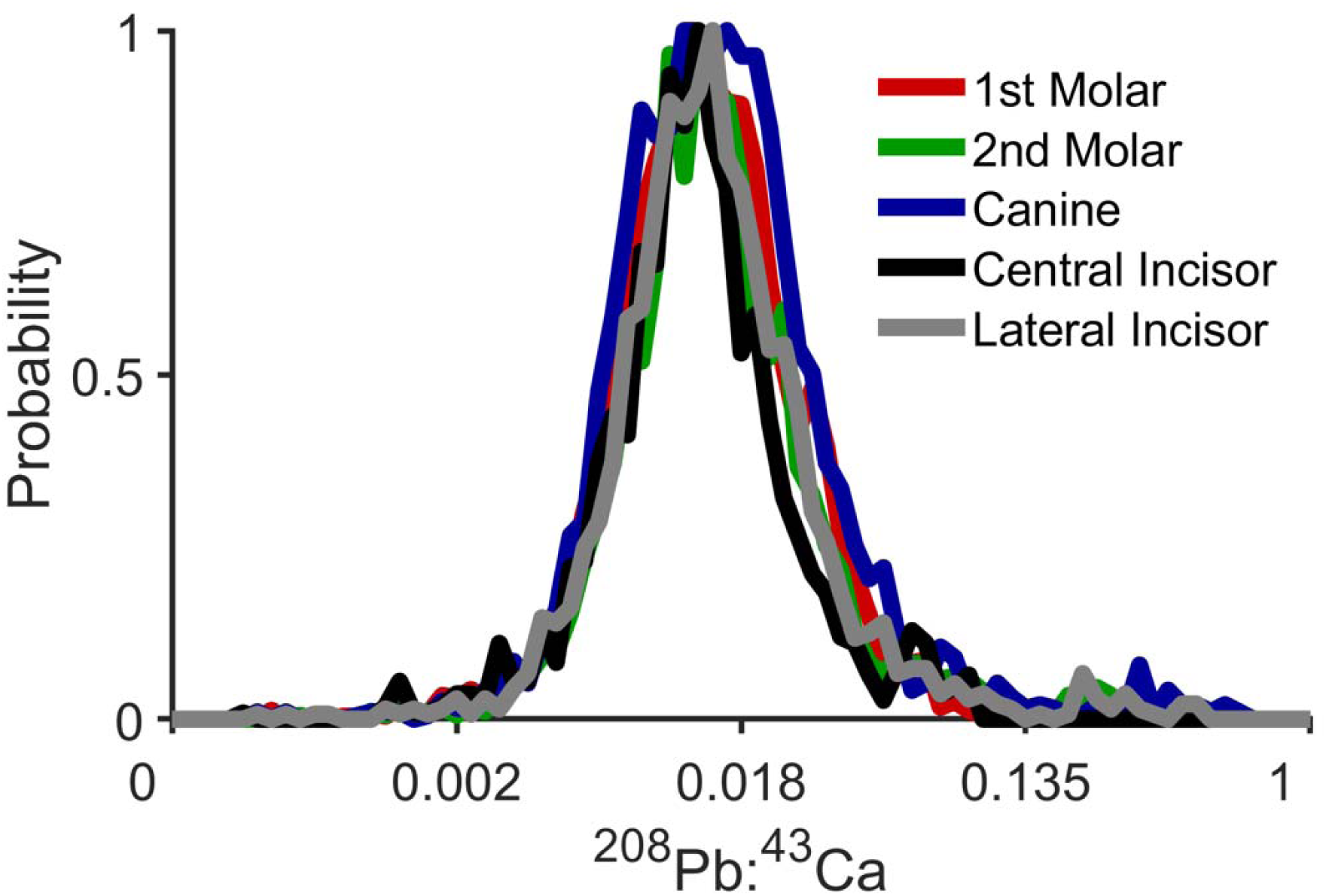
Distribution of tooth-lead levels by tooth type. Tooth-lead levels at individual weeks-since-birth are expressed as relative to calcium levels (i.e., ^208^Pb:^43^Ca). The x-axis is log-transformed. The corresponding distributions are probability histograms, in which frequency counts in each bin were divided by the maximum bin total.

#### Demographics of lead exposure

We first analyzed whether tooth-lead levels varied by annual household income, caregiver education levels, and lead risk. Given the design of the ABCD study, which began collecting data when youth were 9- to 10-years-old, there are no data for income, caregiver education, or lead risk during pregnancy or soon after birth. However, past research has shown, unfortunately, that metrics of socioeconomic status are relatively stable over time,^71^ even across generations.^72,108^ Because we intended to control for these SES-related factors when analyzing neuroanatomical, cognitive, and behavioral outcomes, we evaluated the extent to which tooth-lead levels varied by these factors. For these analyses, results were considered statistically significant at an alpha level of .05.

Due to a high degree of correlation between household income and caregiver education in our sample, *r* = .57, *p* < .001, but lower degrees of correlation between lead risk and both household income, *r* = -0.12, *p* = .011, and caregiver education, *r* = -0.16, *p* = .001, we separately regressed weekly tooth-lead levels on household income and caregiver education. In these analyses of prenatal and postnatal tooth-lead levels (i.e., lead-exposure profiles), we controlled for sex-at-birth and lead risk, along with random intercepts of participant and study site, in 4 sets of models (2 models per set). Here, we employed linear mixed-effects models given the relative monotonicity in how tooth-lead levels changed by WSB (see Figure 1):

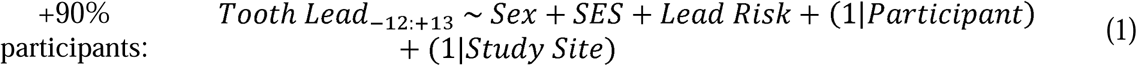

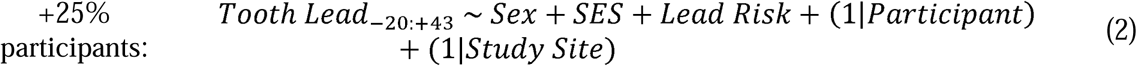

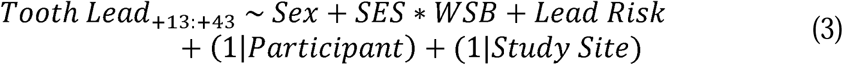

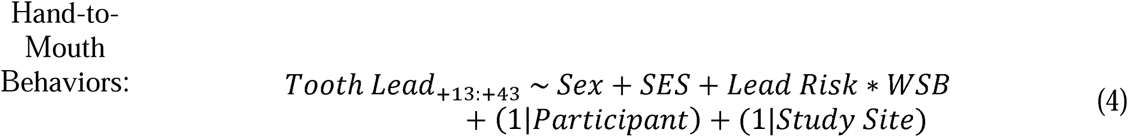

For each model, the subscript following *Tooth Lead* refers to the WSBs analyzed, and *SES* refers to either household income or caregiver education, both centered for their separate analyses. Graphically, household income was partitioned into three brackets: low (<$50,000/year), medium ($50,000-$100,000/year), and high (>$100,000/year). Similarly, caregiver education was split into those with and without a bachelor’s degree, and lead risk was split into three groups: low (lead risk ≤ 3), medium (lead risk = 4-7), and high (lead risk ≥ 8). Subsequent analyses were conducted on cross-tabulated, partitioned subgroups of household income, caregiver education, and lead risk (e.g., low income + no bachelor’s degree + high lead risk vs. high income + bachelor’s degree + low lead risk).

While our primary analyses involved WSBs -12 to +13 to capture when at least 90% of participants had data (Eq. 1), we also expanded this analysis to when at least 25% of participants had data, providing further insight into lead-exposure profiles (Eq. 2). Next, we analyzed whether household income, caregiver education, or lead risk moderated how tooth-lead levels changed postnatally (Eqs. 3-4). Specifically, past research has shown that low SES is associated with the presence of lead-based-paint hazards,^39,109^ which would put individuals living in lower-SES households at greater risk for nondietary lead ingestion.^110,111^ In consideration of the increasing emergence of hand-to-mouth behaviors [starting between 3 and 4 months of age (i.e., 13 to 17 weeks)],^53,55,112^ a primary vehicle of lead exposure in children,^52–54^ we analyzed whether tooth-lead levels increased more rapidly in those living in lower SES households and/or at higher lead risk, from +13 to +43 WSB (Eqs. 3-4). Lastly, we performed simple, descriptive Pearson correlation analyses between individuals’ mean tooth-lead levels (between WSBs -12 to +13) and lead risk plus its housing-age and poverty-rate components. Graphically, for comparison of mean tooth-lead levels to lead risk, we assigned each individual a 1-10 mean tooth-lead level category, in which all individuals were sorted and partitioned into 10 equivalently sized groups (*n*s = 44-45/group).

#### Neuroanatomical structure

We next evaluated associations between tooth-lead levels (-12 to +13 WSB) and cortical structure (thickness, in mm; surface area, in mm^2^) at ABCD’s baseline appointment. Cortical thickness and surface area were analyzed at the regional level (i.e., 68 bilateral cortical regions). These analyses included 437 participants (of 448) whose T_1_-weighted sMRI data passed quality-control inspection, with a total of 11,220 tooth-lead observations (i.e., ∼25.7 tooth-lead measurements per participant). The average age at baseline neuroimaging of these 437 participants was 9.9 years [range: 9.0-11.0 years]. The cortical-thickness and cortical-surface-area general linear mixed-effects models for each of 68 bilateral regions were as follows:

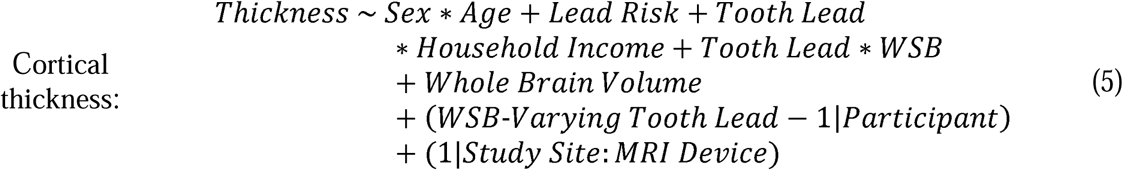

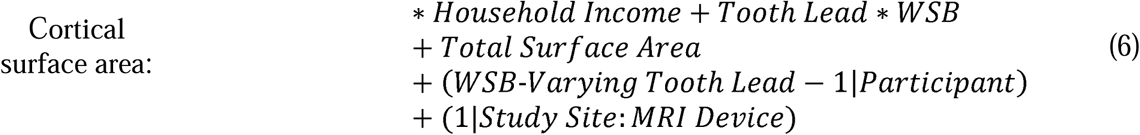

*Sex* was an effects-coded, categorical factor, with male/female sex-at-birth coded as - 1/+1. *Age*, *Tooth Lead*, *Whole Brain Volume*, and *Total Surface Area* were mean-centered, continuous factors. *Lead Risk* and *Household Income* were continuous factors, both centered by 5.5 (i.e., the mean of a 1-10 scale). *WSB* was a continuous factor. (*WSB-Varying Tooth Lead-1*) was operationally defined as participants’ tooth-lead levels by WSB (Tooth Lead:WSB interaction in and of itself). *Study Site* and *MRI Device* were categorical factors. All models used a random initial value for iterative optimization.

*Sex*Age* was included in analyses because of differentially-timed trajectories of cortical development by sex-at-birth.^113^ *Lead Risk* was included in analysis to control for differences in cortical structure for those living in census tracts with varying degrees of lead risk.^45^ *Tooth Lead*Household Income* was included in analyses given our research showing SES-specific relationships between lead risk and whole-brain cortical structure^45^ and to determine if *Household Income* moderated relationships between *Tooth Lead* and cortical structure. *Tooth Lead*WSB* was included in analyses to determine if tooth-lead levels at different timepoints were differentially associated with cortical structure; this interaction (without its corresponding main effects) was allowed to vary by participant to account for individual differences in tooth-lead levels without overfitting the individuals’ single data points in the model (i.e., if we had controlled for these effects at the participant level). *MRI Device* was nested within *Study Site*, as some ABCD study sites use multiple scanners. *Whole Brain Volume* and *Total Surface Area* were controlled for in cortical-thickness and cortical-surface-area models, respectively, given past research showing these whole-brain variables to be more optimal covariates for their corresponding regional analyses.^113^

Two sets of sensitivity analyses were performed. First, for each bilateral region, we computed the Cook’s distance for each tooth-lead – cortical structure datapoint per a simple regression analysis (*Brain Structure ∼ Tooth Lead Level*) to identify multivariate outliers (at the datapoint level). Specifically, datapoints in which their Cook’s distance exceeded 3 times the mean (across datapoints for that analysis) of the Cook’s distance were identified as more influential datapoints (i.e., multivariate outliers) and excluded. Here, between 6 (1.4%) and 21 participants (4.8%) (i.e., 577 [5.1%] to 1,015 observations [9.0%]) were removed from cortical-thickness analyses; between 7 (1.6%) and 18 participants (4.1%) (i.e., 600 [5.3%] to 997 observations [8.9%]) were removed from cortical-surface-area analyses. Second, sensitivity analyses were performed in which the same models as above (Eqs. 5-6) were run separately for prenatal (WSBs: [-12, 0]; 5,567 observations across 437 participants) and postnatal lead exposure (WSBs: [+1, +13]; 5,653 observations across 437 participants).

Effect sizes were represented by partial correlation coefficients (*r_p_*), which control for all model covariates and are calculated using the corresponding *t*-statistic and degrees of freedom.^114^ The 95% confidence intervals (CIs) of the effect sizes were derived from the sample variance of the partial correlation.^115^ Given the number of brain regions analyzed, the Benjamini-Hochberg^116^ false-discovery-rate (FDR) algorithm was used to correct for multiple comparisons by metric, separately, within each set of analyses (e.g., prenatal tooth-lead levels as a predictor for cortical thickness), relative to a *p*-value threshold of .005.^51^ Model output for cortical analyses is shown in Tables S12-S91.

#### Cognition and behavior

As with the neuroanatomical analyses, we performed general linear mixed-effects models to evaluate associations between tooth-lead levels (WSBs: [-12, +13]) and (1) cognitive performance on the NIH Toolbox and (2) caregiver-reported behavior via the CBCL. The corresponding models were identical to Eqs. 5-6 with two exceptions: (1) We did not control for a whole-brain variable (i.e., whole-brain volume or total surface area), and (2) we did not control for MRI device in the random-effects terms (but did control for ABCD study site). An additional difference was that more participants had valid NIH Toolbox and CBCL data; 440 and 444 participants had NIH Toolbox composite scores for fluid and crystallized cognition, respectively (i.e., 440 participants had a total composite cognitive score). All 448 participants had CBCL data. The average age of these 448 participants was 9.9 years [range: 9.0-11.0 years; SD = 0.6]. For the NIH Toolbox, we analyzed uncorrected scores, in line with our past research^45^; for the CBCL, standardized scores to more closely approximate a normal distribution and for their clinical utility.

First, we analyzed the total cognitive score and total problem behaviors from the NIH Toolbox and CBCL, respectively. Significant effects of tooth-lead levels on each (*p*s < .005) led us to evaluate whether their respective components were differentially associated with tooth-lead levels (NIH Toolbox: fluid cognition, crystallized cognition; CBCL: externalizing behavior, internalizing behavior). A significant effect of tooth-lead levels on crystallized cognition (*p* < .005) led us to determine whether performances on the Picture Vocabulary or Oral Reading tests were differentially associated with tooth-lead levels. At the final stage of these reductionist analyses, we also (1) determined whether household income (SES) moderated the relationship between tooth-lead levels and the corresponding outcome and (2) performed sensitivity analyses for prenatal lead exposure (WSBs: [-12, 0];), postnatal lead exposure (WSBs: [+1, +13];), and excluding individual datapoints with larger Cook’s distances values (i.e., multivariate outliers) per simple-regression analyses. For the latter, 11 (2.5%) (i.e., 837 [7.3%] observations) were removed from the corresponding NIH-Toolbox analysis; 16 (3.6%) (i.e., 751 [6.5%] observations) were removed from the corresponding CBCL analysis. As above, effect sizes were represented by partial correlation coefficients (*r_p_*), and the *p*-value threshold was .005.^51^ Model output for cortical analyses is shown in Tables S92-S105. For visual display (to convey any interactions involving lead exposure within the NIH Toolbox and CBCL analyses), we split participants (based on their mean tooth-lead levels) into three groups [i.e., bottom-third (<33%ile), middle-third (33-66%ile), and top-third (≥67%ile)].

## Supporting information

Supplemental Figures and Tables (Part 1)

Supplemental Tables (Part 2)

## Data Availability

ABCD data are publicly available through the NIH Brain Development Cohorts Data Hub (https://www.nbdc-datahub.org/).

## Acknowledgments and Funding

Data used in the preparation of this article were obtained from the Adolescent Brain Cognitive Development™ (ABCD) Study (abcdstudy.org), held in the NIH Brain Development Cohorts Data Sharing Platform (https://www.nbdc-datahub.org/). This is a multisite, longitudinal study designed to recruit more than 10,000 children aged 9–10 and follow them over 10 years into early adulthood. The ABCD Study® is supported by the National Institutes of Health and additional federal partners under award numbers: U01DA041048, U01DA050989, U01DA051016, U01DA041022, U01DA051018, U01DA051037, U01DA050987, U01DA041174, U01DA041106, U01DA041117, U01DA041028, U01DA041134, U01DA050988, U01DA051039, U01DA041156, U01DA041025, U01DA041120, U01DA051038, U01DA041148, U01DA041093, U01DA041089, U24DA041123, U24DA041147. A full list of supporters is available at Federal Partners – ABCD Study (https://abcdstudy.org/about/federal-partners/). Additional support for this work was made possible from R01-ES032295, R01-ES031074, R01-ES031247 (Sowell), and K01-DK135847 (Adise), P30-ES007048 (McConnell), and 5P2CES033433 (McConnell). ABCD Consortium investigators designed and implemented the study and/or provided data but did not necessarily participate in the analysis or writing of this report. This manuscript reflects the views of the authors and may not reflect the opinions or views of the NIH or ABCD Consortium investigators. The ABCD data repository grows and changes over time. The ABCD data used in this report came from ABCD Public Data Release 7.0 (https://doi.org/10.82525/8f3w-5260).

## Author Contributions

A.T.M., S.A., R.M., V.M., M.A., and E.R.S conceived and designed the experiments/analysis. A.T.M., E.K., M.M., and M.A. collected the data. A.T.M., S.A., M.K., M.M., V.M., and M.A. contributed data or analysis tools. A.T.M., E.K., M.K., M.M., and M.A. analyzed the data. A.T.M., M.M., M.A., and E.R.S. wrote the paper.

## Materials & Correspondence

Correspondence to Elizabeth R. Sowell.

