## Supplemental Figures and Tables (Part 1) for "Dentine markers of pre/early postnatal lead exposure links with brain, cognitive, and behavioral outcomes in adolescents"

**Supplemental Material**

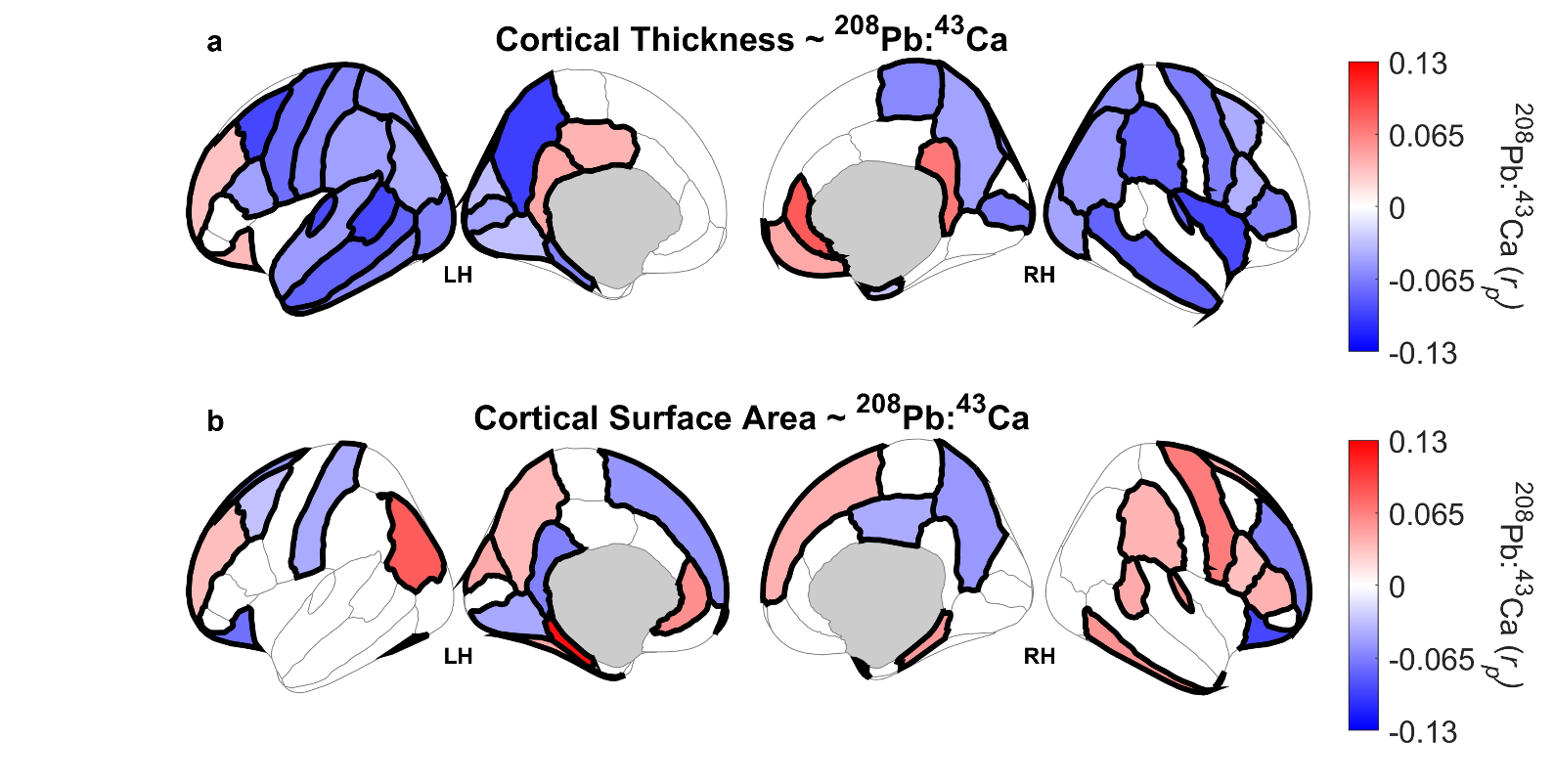

**Figure S3. Associations between tooth-lead (Pb) levels and cortical structure, excluding datapoints designated as multivariate outliers.** Bilateral cortical regions are color-coded based on the strength of the association between tooth-lead levels and cortical structure, controlling for sex, age, week-since-birth (WSB), household income, lead-exposure risk, Adolescent Brain Cognitive Development (ABCD) study site and magnetic resonance imaging scanner, and overall brain size (cortical thickness: whole-brain volume; cortical surface area: total surface area). For each bilateral region, we computed the Cook’s distance for each tooth-lead – cortical structure datapoint per a simple regression analysis (*Brain Structure ~ Tooth Lead Level*) to identify multivariate outliers. Datapoints in which their Cook’s distance exceeded 3 times the mean (across datapoints for that analysis) of the Cook’s distance were identified as multivariate outliers and excluded. Color-coding (per partial correlation coefficients, *r_p_*) is specific to analyses including weekly tooth-lead levels between ‑12 and +13 WSB, in which all non-excluded tooth-lead levels within that 26-WSB range were included in analysis. Regions are shaded if those associations passed false-discovery-rate (FDR) correction; non-shaded regions indicate that the corresponding association did not pass FDR correction. Regions with a red (blue) shading indicate positive (negative) relationships between tooth-lead levels and cortical structure. Tooth-lead levels were expressed relative to calcium (Ca) content in each tooth (^208^Pb:^43^Ca ratio). LH = left hemisphere. RH = right hemisphere.

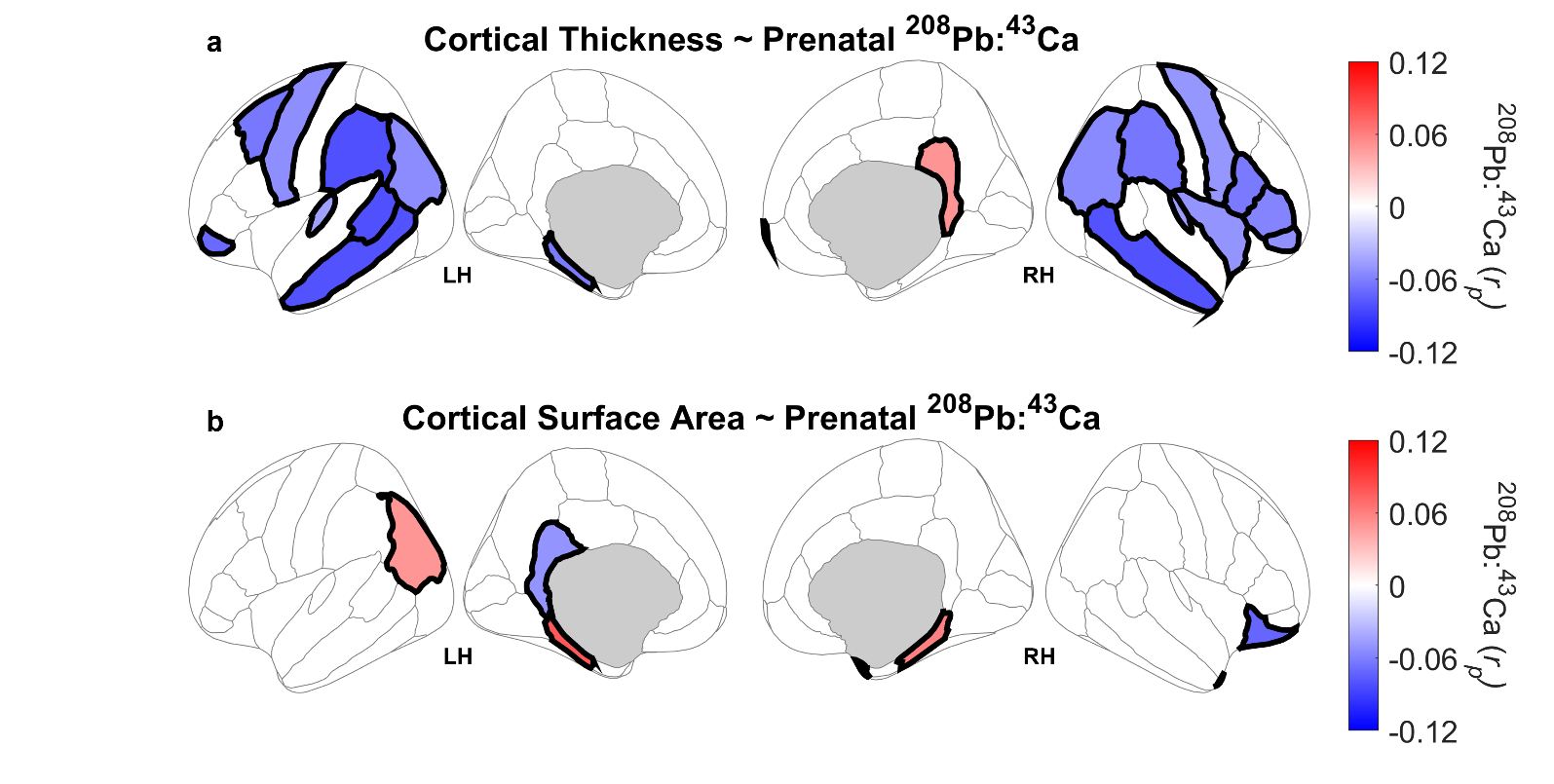

**Figure S4. Associations between prenatal tooth-lead (Pb) levels and cortical structure.** Bilateral cortical regions are color-coded based on the strength of the association between prenatal tooth-lead levels and cortical structure, controlling for sex, age, week-since-birth (WSB), household income, lead-exposure risk, Adolescent Brain Cognitive Development (ABCD) study site and magnetic resonance imaging scanner, and overall brain size (cortical thickness: whole-brain volume; cortical surface area: total surface area). Color-coding (per partial correlation coefficients, *r_p_*) is specific to analyses including weekly tooth-Pb levels between ‑12 and 0 WSB, in which all tooth-lead levels within that 13-WSB range were included in analysis. Regions are shaded if those associations passed false-discovery-rate (FDR) correction; non-shaded regions indicate that the corresponding association did not pass FDR correction. Regions with a red (blue) shading indicate positive (negative) relationships between tooth-lead levels and cortical structure. Tooth-lead levels were expressed relative to calcium (Ca) content in each tooth (^208^Pb:^43^Ca ratio). LH = left hemisphere. RH = right hemisphere.

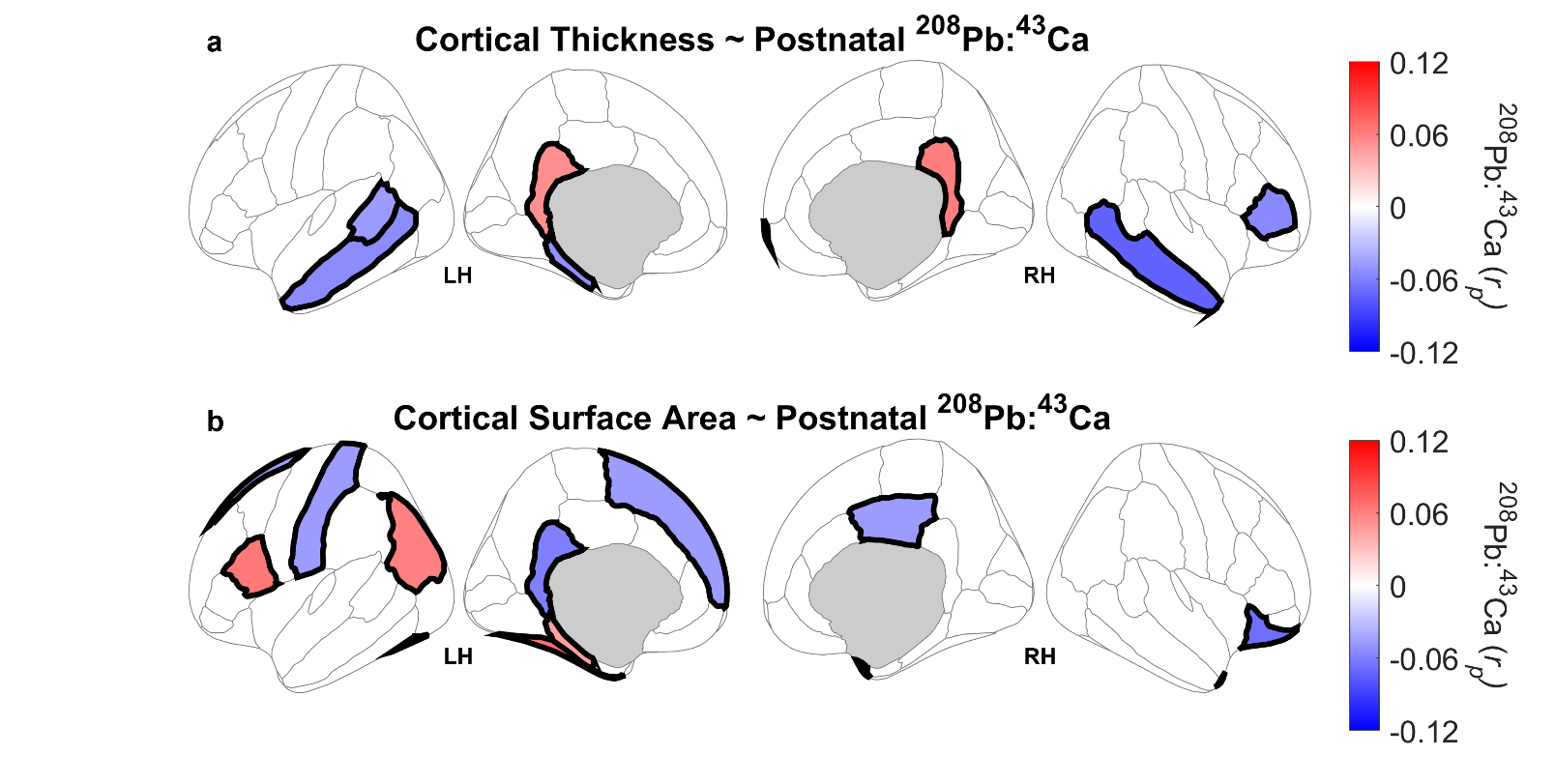

**Figure S5. Associations between postnatal tooth-lead (Pb) levels and cortical structure.** Bilateral cortical regions are color-coded based on the strength of the association between postnatal tooth-lead levels and cortical structure, controlling for sex, age, week-since-birth (WSB), household income, lead-exposure risk, Adolescent Brain Cognitive Development (ABCD) study site and magnetic resonance imaging scanner, and overall brain size (cortical thickness: whole-brain volume; cortical surface area: total surface area). Color-coding (per partial correlation coefficients, *r_p_*) is specific to analyses including weekly tooth-Pb levels between +1 and +13 WSB, in which all tooth-lead levels within that 13-WSB range were included in analysis. Regions are shaded if those associations passed false-discovery-rate (FDR) correction; non-shaded regions indicate that the corresponding association did not pass FDR correction. Regions with a red (blue) shading indicate positive (negative) relationships between tooth-lead levels and cortical structure. Tooth-lead levels were expressed relative to calcium (Ca) content in each tooth (^208^Pb:^43^Ca ratio). LH = left hemisphere. RH = right hemisphere.

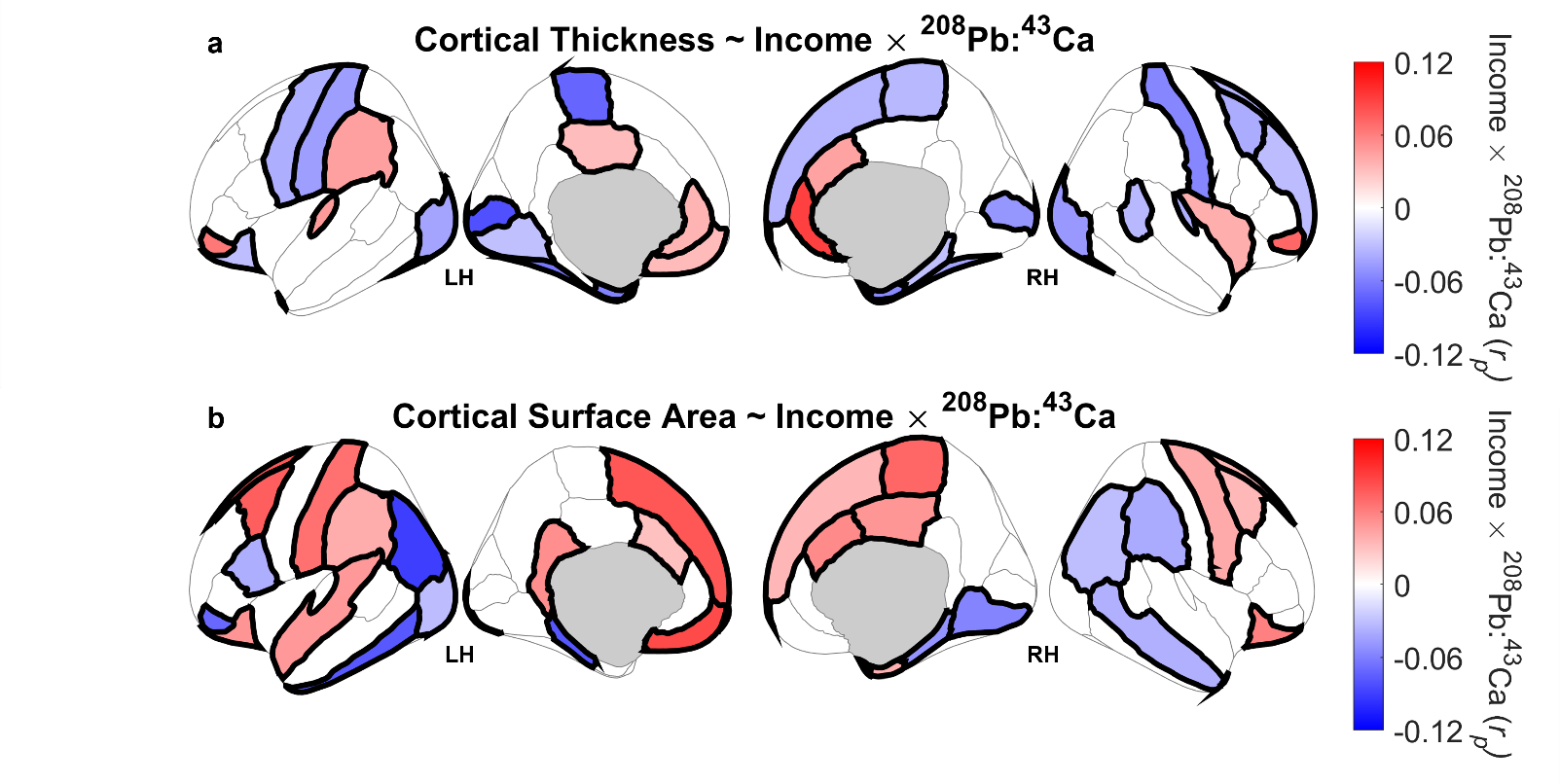

**Figure S6. Household Income × Tooth-Lead (Pb) Level interactions on cortical structure.** Bilateral cortical regions are color-coded based on the strength of the interaction between household income and tooth-lead levels on cortical structure, controlling for sex, age, week-since-birth (WSB), household income, lead-exposure risk, Adolescent Brain Cognitive Development (ABCD) study site and magnetic resonance imaging scanner, and overall brain size (cortical thickness: whole-brain volume; cortical surface area: total surface area). Color-coding (per partial correlation coefficients, *r_p_*) is specific to analyses including weekly tooth-lead levels between ‑12 and +13 WSB, in which all tooth-lead levels within that 26-WSB range were included in analysis. Regions are shaded if those interactions passed false-discovery-rate (FDR) correction; non-shaded regions indicate that the corresponding interaction did not pass FDR correction. Tooth-Pb levels were expressed relative to calcium (Ca) content in each tooth (^208^Pb:^43^Ca ratio). LH = left hemisphere. RH = right hemisphere.

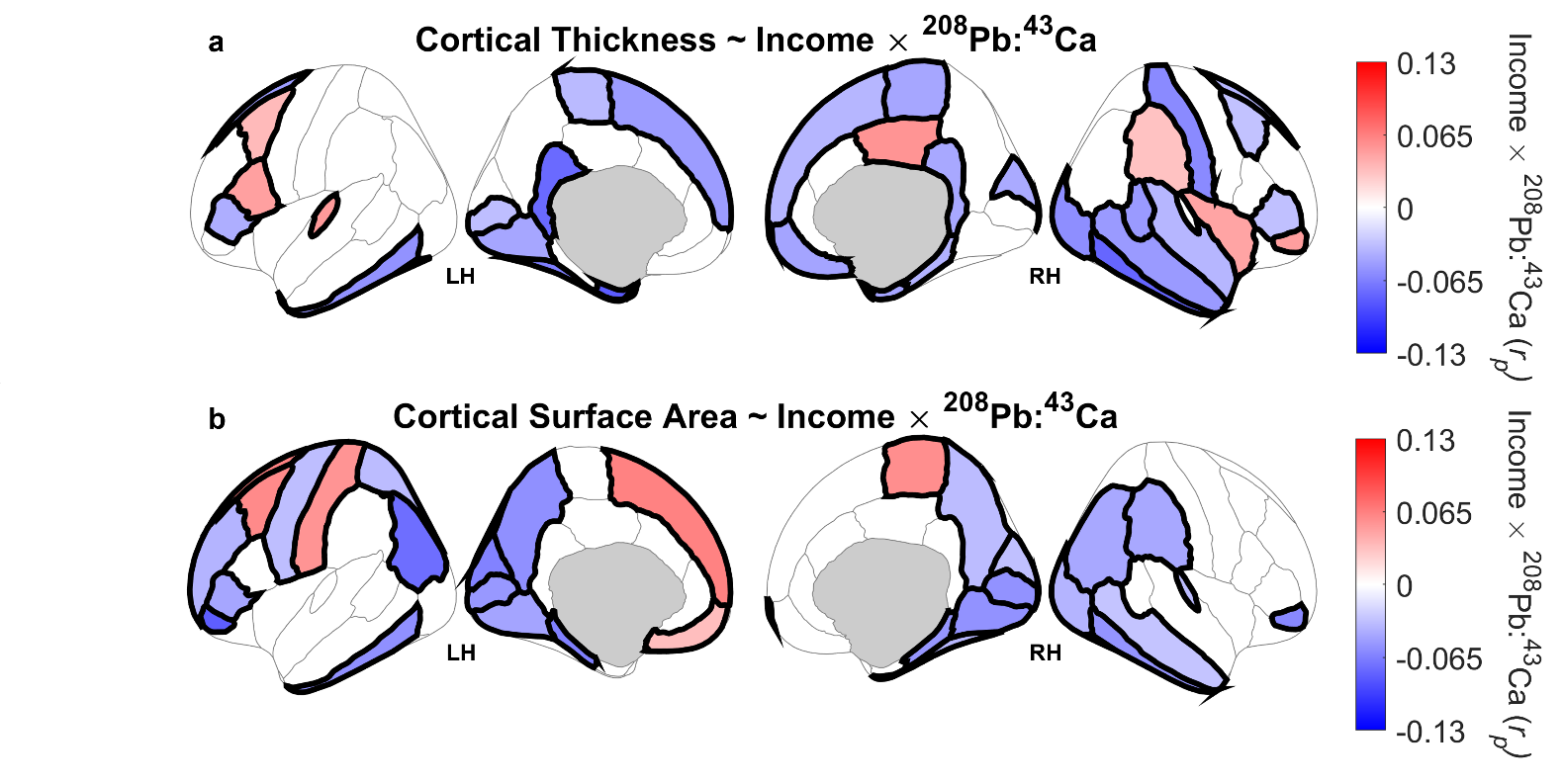

**Figure S7. Household Income × Tooth-Lead (Pb) Level interactions on cortical structure, excluding datapoints designated as multivariate outliers.** Bilateral cortical regions are color-coded based on the strength of the interaction between household income and tooth-lead levels on cortical structure, controlling for sex, age, week-since-birth (WSB), household income, lead-exposure risk, Adolescent Brain Cognitive Development (ABCD) study site and magnetic resonance imaging scanner, and overall brain size (cortical thickness: whole-brain volume; cortical surface area: total surface area). For each bilateral region, we computed the Cook’s distance for each tooth-lead – cortical structure datapoint per a simple regression analysis (*Brain Structure ~ Tooth Lead Level*) to identify multivariate outliers. Datapoints in which their Cook’s distance exceeded 3 times the mean (across datapoints for that analysis) of the Cook’s distance were identified as multivariate outliers and excluded. Color-coding (per partial correlation coefficients, *r_p_*) is specific to analyses including weekly tooth-lead levels between ‑12 and +13 WSB, in which all non-excluded tooth-lead levels within that 26-WSB range were included in analysis. Regions are shaded if those interactions passed false-discovery-rate (FDR) correction; non-shaded regions indicate that the corresponding interaction did not pass FDR correction. Tooth-Pb levels were expressed relative to calcium (Ca) content in each tooth (^208^Pb:^43^Ca ratio). LH = left hemisphere. RH = right hemisphere.

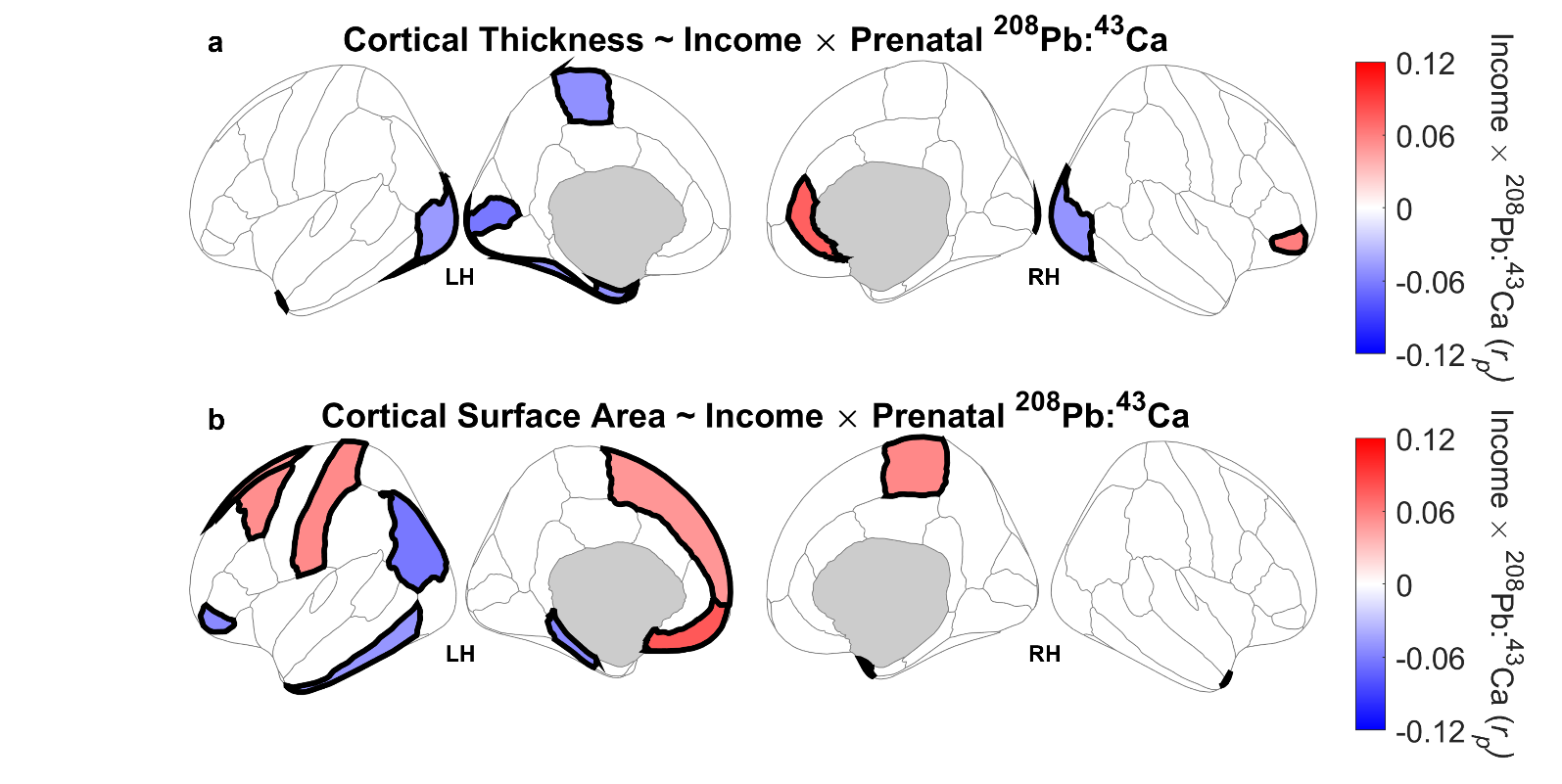

**Figure S8. Household Income × Prenatal Tooth-Lead (Pb) Level interactions on cortical structure.** Bilateral cortical regions are color-coded based on the strength of the interaction between household income and prenatal tooth-lead levels on cortical structure, controlling for sex, age, week-since-birth (WSB), household income, lead-exposure risk, Adolescent Brain Cognitive Development (ABCD) study site and magnetic resonance imaging scanner, and overall brain size (cortical thickness: whole-brain volume; cortical surface area: total surface area). Color-coding (per partial correlation coefficients, *r_p_*) is specific to analyses including weekly tooth-lead levels between ‑12 and 0 WSB, in which all tooth-lead levels within that 13-WSB range were included in analysis. Regions are shaded if those interactions passed false-discovery-rate (FDR) correction; non-shaded regions indicate that the corresponding interaction did not pass FDR correction. Tooth-Pb levels were expressed relative to calcium (Ca) content in each tooth (^208^Pb:^43^Ca ratio). LH = left hemisphere. RH = right hemisphere.

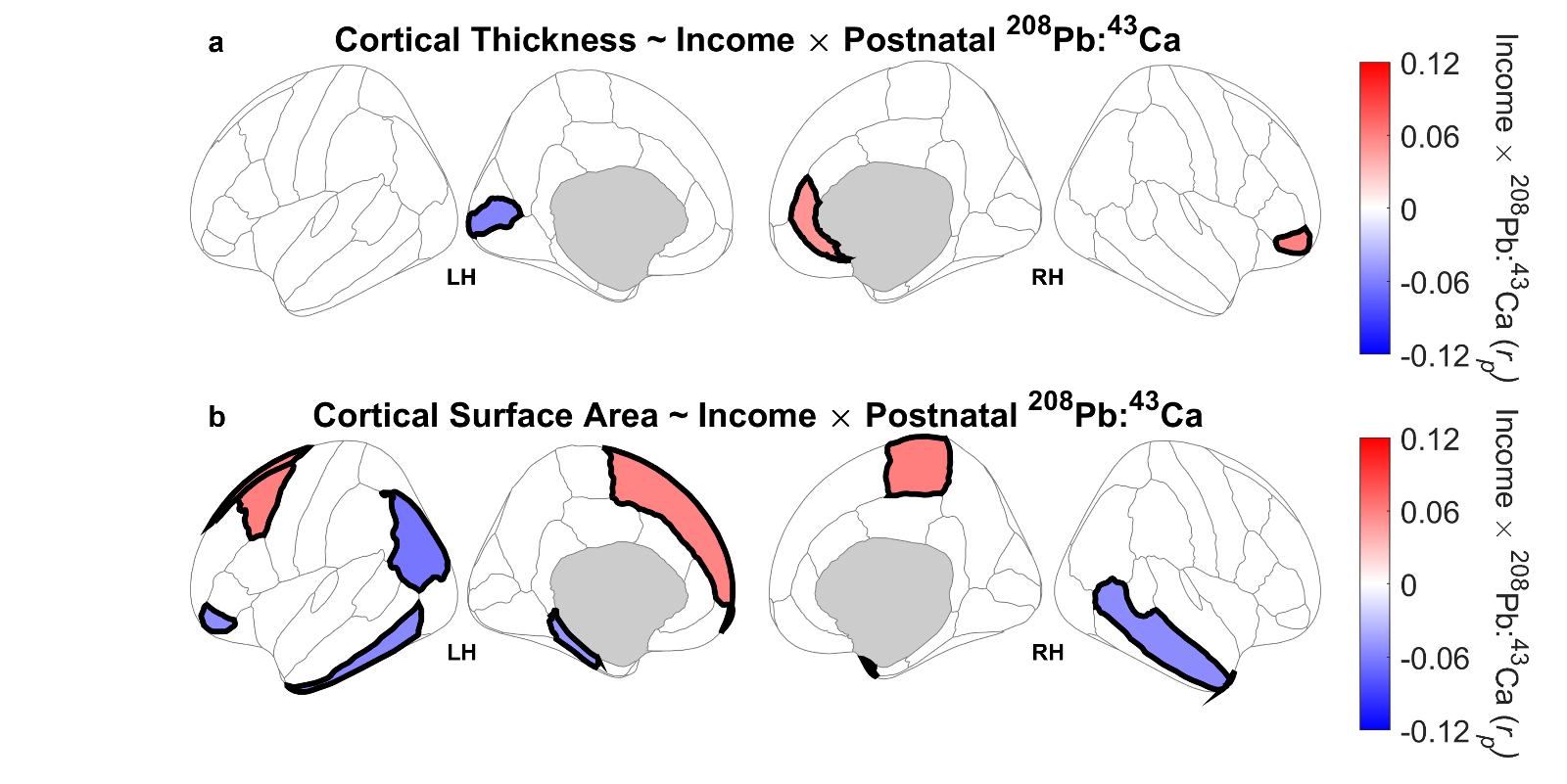

**Figure S9. Household Income × Postnatal Tooth-Lead (Pb) Level interactions on cortical structure.** Bilateral cortical regions are color-coded based on the strength of the interaction between household income and postnatal tooth-lead levels on cortical structure, controlling for sex, age, week-since-birth (WSB), household income, lead-exposure risk, Adolescent Brain Cognitive Development (ABCD) study site and magnetic resonance imaging scanner, and overall brain size (cortical thickness: whole-brain volume; cortical surface area: total surface area). Color-coding (per partial correlation coefficients, *r_p_*) is specific to analyses including weekly tooth-lead levels between +1 and +13 WSB, in which all tooth-lead levels within that 13-WSB range were included in analysis. Regions are shaded if those interactions passed false-discovery-rate (FDR) correction; non-shaded regions indicate that the corresponding interaction did not pass FDR correction. Tooth-Pb levels were expressed relative to calcium (Ca) content in each tooth (^208^Pb:^43^Ca ratio). LH = left hemisphere. RH = right hemisphere.

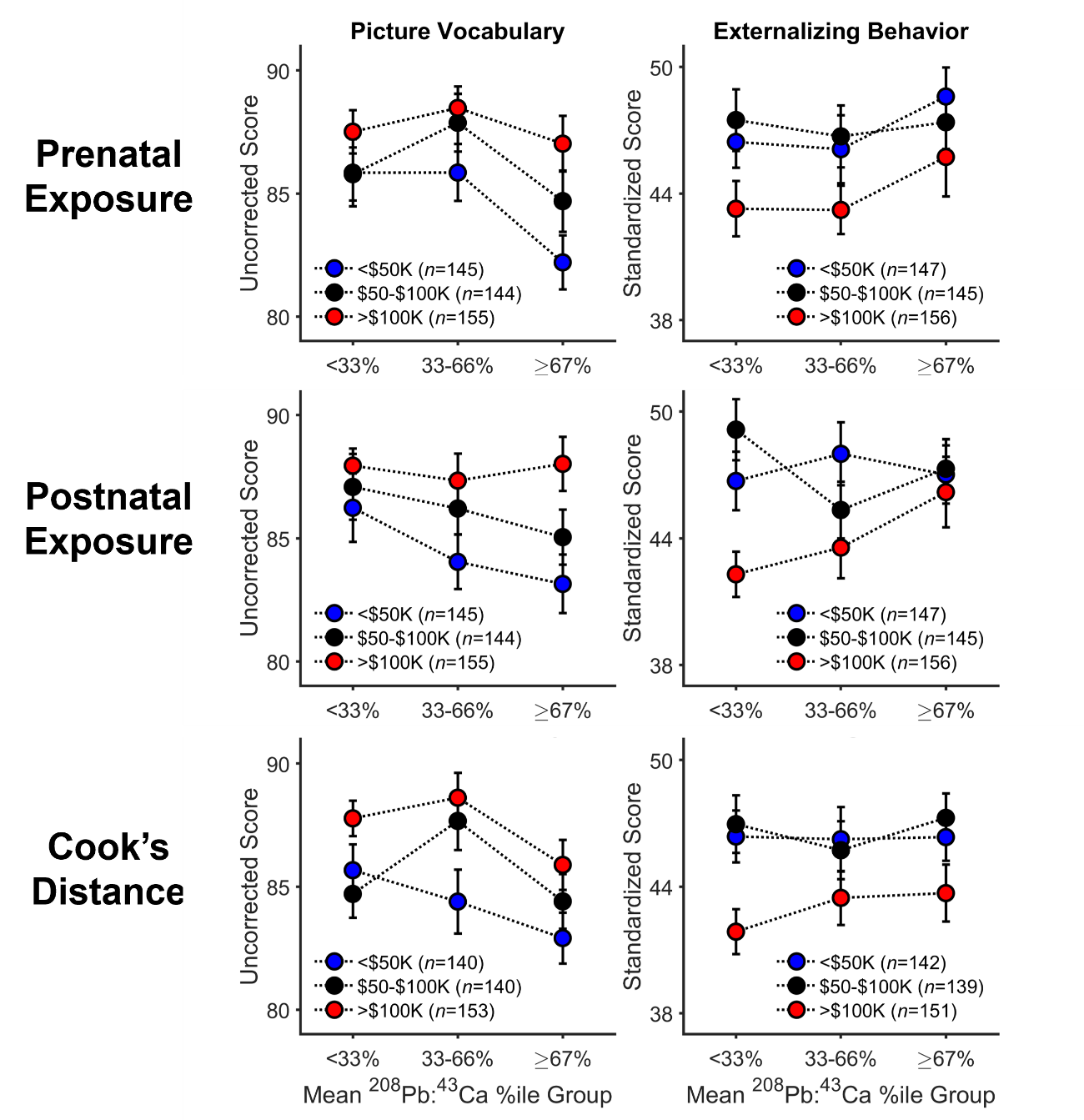

**Figure S10. Household Income × Tooth-Lead (Pb) Level interactions on picture vocabulary and externalizing behavior**. Lead exposure was expressed relative to calcium (Ca) content in each tooth (^208^Pb:^43^Ca ratio); for ease of interpretation, participants’ mean tooth-lead levels [top row: from ‑12 to 0 weeks since birth (WSB); middle row: from +1 to +13 WSB; bottom row: from ‑12 to +13 WSB, excluding datapoints designated as multivariate outliers] were collapsed into 3 groups (i.e., the bottom third, middle third, and top third of mean tooth-lead levels). Within each panel, data are also split based on caregiver-reported annual household income, with the total numbers of participants in each subgroup shown. The corresponding analyses included individual tooth-lead levels by WSB and also controlled for sex, age, lead-exposure risk, and Adolescent Brain Cognitive Development (ABCD) study site. Each datapoint reflects the mean ± 1 standard error of the mean (SEM).

**Table S2. Linear mixed-effects model output for the analysis of tooth-lead levels from ‑20 to +43 weeks since birth (WSB), with sex, maximum caregiver education, and risk of Pb exposure as predictors.**

|  | *t*(21451) | *p* | *b* | 95% CI |
| --- | --- | --- | --- | --- |
| Intercept | -80.84 | < .001 | -4.00 | [-4.10, -3.90] |
| Sex | -1.31 | .190 | -0.04 | [-0.09, 0.02] |
| Maximum Caregiver Education | -2.57 | .010 | -0.07 | [-0.13, -0.02] |
| Risk of Lead Exposure | 2.98 | .003 | 0.03 | [0.01, 0.05] |

**Note**: The linear mixed-effects model incorporates testing the statistical significance of coefficients against a *t*-distribution. Sex was a categorical factor, effect coded with Male/Female as ‑1/+1. Maximum caregiver education and risk of lead exposure were centered, continuous factors. The random-effects structure included separate random intercepts for participant ID number and for the participant’s study site. Tooth-lead levels were operationally defined relative to calcium (Ca) content in each tooth (^208^Pb:^43^Ca ratio) to control for overall mineral content. Analysis included 21,455 data points. The model accounted for 61.3% of the variance in the data (*R*^2^ = .613, adjusted *R*^2^ = .613).

**Table S3. Linear mixed-effects model output for the analysis of tooth-lead levels from ‑20 to +43 weeks since birth (WSB), with sex, annual household income, and risk of Pb exposure as predictors.**

|  | *t*(21451) | *p* | *b* | 95% CI |
| --- | --- | --- | --- | --- |
| Intercept | -84.81 | < .001 | -4.02 | [-4.12, -3.93] |
| Sex | -1.34 | .182 | -0.04 | [-0.10, 0.02] |
| Household Income | -2.01 | .044 | -0.03 | [-0.06, -0.001] |
| Risk of Lead Exposure | 3.10 | .002 | 0.03 | [0.01, 0.05] |

**Note**: The linear mixed-effects model incorporates testing the statistical significance of coefficients against a *t*-distribution. Sex was a categorical factor, effect coded with Male/Female as ‑1/+1. Household income and risk of lead exposure were centered, continuous factors. The random-effects structure included separate random intercepts for participant ID number and for the participant’s study site. Tooth-lead levels were operationally defined relative to calcium (Ca) content in each tooth (^208^Pb:^43^Ca ratio) to control for overall mineral content. Analysis included 21,455 data points. The model accounted for 61.3% of the variance in the data (*R*^2^ = .613, adjusted *R*^2^ = .613).

**Table S4. Linear mixed-effects model output for the analysis of tooth-lead levels from ‑12 to +13 weeks since birth (WSB), with sex, maximum caregiver education, and risk of Pb exposure as predictors.**

|  | *t*(11498) | *p* | *b* | 95% CI |
| --- | --- | --- | --- | --- |
| Intercept | -79.05 | < .001 | -4.11 | [-4.21, -4.01] |
| Sex | -1.57 | .116 | -0.05 | [-0.10, 0.01] |
| Maximum Caregiver Education | -2.34 | .019 | -0.07 | [-0.12, -0.01] |
| Risk of Lead Exposure | 2.66 | .008 | 0.03 | [0.01, 0.05] |

**Note**: The linear mixed-effects model incorporates testing the statistical significance of coefficients against a *t*-distribution. Sex was a categorical factor, effect coded with Male/Female as ‑1/+1. Maximum caregiver education and risk of lead exposure were centered, continuous factors. The random-effects structure included separate random intercepts for participant ID number and for the participant’s study site. Tooth-lead levels were operationally defined relative to calcium (Ca) content in each tooth (^208^Pb:^43^Ca ratio) to control for overall mineral content. Analysis included 11,502 data points. The model accounted for 77.2% of the variance in the data (*R*^2^ = .772, adjusted *R*^2^ = .772).

**Table S5. Linear mixed-effects model output for the analysis of tooth-lead levels from ‑12 to +13 weeks since birth (WSB), with sex, annual household income, and risk of Pb exposure as predictors.**

|  | *t*(11498) | *p* | *b* | 95% CI |
| --- | --- | --- | --- | --- |
| Intercept | -82.36 | < .001 | -4.13 | [-4.23, -4.03] |
| Sex | -1.60 | .109 | -0.05 | [-0.10, 0.01] |
| Household Income | -2.10 | .036 | -0.03 | [-0.06, -0.002] |
| Risk of Lead Exposure | 2.74 | .006 | 0.03 | [0.01, 0.05] |

**Note**: The linear mixed-effects model incorporates testing the statistical significance of coefficients against a *t*-distribution. Sex was a categorical factor, effect coded with Male/Female as ‑1/+1. Household income and risk of lead exposure were centered, continuous factors. The random-effects structure included separate random intercepts for participant ID number and for the participant’s study site. Tooth-lead levels were operationally defined relative to calcium (Ca) content in each tooth (^208^Pb:^43^Ca ratio) to control for overall mineral content. Analysis included 11,502 data points. The model accounted for 77.2% of the variance in the data (*R*^2^ = .772, adjusted *R*^2^ = .772).

**Table S6. Linear mixed-effects model output for the analysis of tooth-lead levels from -20 to +43 weeks since birth (WSB), with sex and an aggregate Pb-risk category as predictors.**

|  | *t*(4482) | *p* | *b* | 95% CI |
| --- | --- | --- | --- | --- |
| Intercept | -72.84 | < .001 | -3.98 | [-4.09, -3.87] |
| Sex | -2.54 | .011 | -0.13 | [-0.24, -0.03] |
| Lead-Risk Category | 4.62 | < .001 | 0.25 | [0.15, 0.36] |

**Note**: The linear mixed-effects model incorporates testing the statistical significance of coefficients against a *t*-distribution. Sex was a categorical factor, effect coded with Male/Female as ‑1/+1. Lead-Risk Category was also a categorical factor, effect coded with Low Risk/High Risk as ‑1/+1. “Low Risk” was a cross-sectional category of 58 participants living in households with incomes greater than $100,000/year, with caregiver(s) whose maximum education level was at least a bachelor's degree, and in census tracts with a lead-risk score of 1-3. “High Risk” was a cross-sectional category of 34 participants living in households with incomes less than $50,000/year, with caregiver(s) whose maximum education level was less than a bachelor's degree, and in census tracts with a lead-risk score of 8-10. The random-effects structure included separate random intercepts for participant ID number and for the participant’s study site. Tooth-lead levels were operationally defined relative to calcium (Ca) content in each tooth (^208^Pb:^43^Ca ratio) to control for overall mineral content. Analysis included 4,485 data points. The model accounted for 56.7% of the variance in the data (*R*^2^ = .567, adjusted *R*^2^ = .566).

**Table S7. Linear mixed-effects model output for the analysis of tooth-lead levels from -12 to +13 weeks since birth (WSB), with sex and an aggregate Pb-risk category as predictors.**

|  | *t*(2359) | *p* | *b* | 95% CI |
| --- | --- | --- | --- | --- |
| Intercept | -80.41 | < .001 | -4.11 | [-4.21, -4.01] |
| Sex | -3.14 | .002 | -0.16 | [-0.25, -0.06] |
| Lead-Risk Category | 4.64 | < .001 | 0.24 | [0.14, 0.34] |

**Note**: The linear mixed-effects model incorporates testing the statistical significance of coefficients against a *t*-distribution. Sex was a categorical factor, effect coded with Male/Female as ‑1/+1. Pb-Risk Category was also a categorical factor, effect coded with Low Risk/High Risk as ‑1/+1. “Low Risk” was a cross-sectional category of 58 participants living in households with incomes greater than $100,000/year, with caregiver(s) whose maximum education level was at least a bachelor's degree, and in census tracts with a lead-risk score of 1-3. “High Risk” was a cross-sectional category of 34 participants living in households with incomes less than $50,000/year, with caregiver(s) whose maximum education level was less than a bachelor's degree, and in census tracts with a lead-risk score of 8-10. The random-effects structure included separate random intercepts for participant ID number and for the participant’s study site. Tooth-lead levels were operationally defined relative to calcium (Ca) content in each tooth (^208^Pb:^43^Ca ratio) to control for overall mineral content. Analysis included 2,362 data points. The model accounted for 68.2% of the variance in the data (*R*^2^ = .682, adjusted *R*^2^ = .681).

**Table S8. Linear mixed-effects model output for the analysis of tooth-lead levels from +13 to +43 weeks since birth (WSB), with sex, maximum caregiver education, weeks-since-birth (WSB), risk of Pb exposure, and WSB × Maximum Caregiver Education as predictors.**

|  | *t*(8161) | *p* | *b* | 95% CI |
| --- | --- | --- | --- | --- |
| Intercept | -86.03 | < .001 | -4.65 | [-4.76, -4.55] |
| Weeks-Since-Birth (WSB) | 51.92 | < .001 | 0.04 | [0.03, 0.04] |
| Sex | -1.77 | .077 | -0.06 | [-0.13, 0.01] |
| Maximum Caregiver Education | -0.38 | .706 | -0.01 | [-0.08, 0.06] |
| Risk of Lead Exposure | 2.24 | .025 | 0.03 | [0.003, 0.05] |
| WSB × Maximum Caregiver Education | -2.89 | .004 | -0.001 | [-0.002, -0.0005] |

**Note**: The linear mixed-effects model incorporates testing the statistical significance of coefficients against a *t*-distribution. Sex was a categorical factor, effect coded with Male/Female as ‑1/+1. WSB, maximum caregiver education and risk of lead exposure were centered, continuous factors. The random-effects structure included separate random intercepts for participant ID number and for the participant’s study site. Tooth-lead levels were operationally defined relative to calcium (Ca) content in each tooth (^208^Pb:^43^Ca ratio) to control for overall mineral content. Analysis included 8,167 data points. The model accounted for 82.2% of the variance in the data (*R*^2^ = .822, adjusted *R*^2^ = .822).

**Table S9. Linear mixed-effects model output for the analysis of tooth-lead levels from +13 to +43 weeks since birth (WSB), with sex, household income, weeks-since-birth (WSB), risk of Pb exposure, and WSB × Household Income as predictors.**

|  | *t*(8161) | *p* | *b* | 95% CI |
| --- | --- | --- | --- | --- |
| Intercept | -93.12 | < .001 | -4.71 | [-4.81, -4.61] |
| Weeks-Since-Birth (WSB) | 56.34 | < .001 | 0.04 | [0.04, 0.04] |
| Sex | -1.85 | .065 | -0.06 | [-0.13, 0.004] |
| Household Income | 1.41 | .158 | 0.02 | [-0.01, 0.06] |
| Risk of Lead (Pb) Exposure | 2.26 | .024 | 0.03 | [0.004, 0.05] |
| WSB × Household Income | -8.67 | < .001 | -0.002 | [-0.003, -0.002] |

**Note**: The linear mixed-effects model incorporates testing the statistical significance of coefficients against a *t*-distribution. Sex was a categorical factor, effect coded with Male/Female as ‑1/+1. WSB, household income, and risk of lead exposure were centered, continuous factors. The random-effects structure included separate random intercepts for participant ID number and for the participant’s study site. Tooth-lead levels were operationally defined relative to calcium (Ca) content in each tooth (^208^Pb:^43^Ca ratio) to control for overall mineral content. Analysis included 8,167 data points. The model accounted for 82.4% of the variance in the data (*R*^2^ = .824, adjusted *R*^2^ = .824).

**Table S10. Linear mixed-effects model output for the analysis of tooth-lead levels from +13 to +43 weeks since birth (WSB), with sex, maximum caregiver education, weeks-since-birth (WSB), risk of Pb exposure, and WSB × Risk of Pb Exposure as predictors.**

|  | *t*(8161) | *p* | *b* | 95% CI |
| --- | --- | --- | --- | --- |
| Intercept | -86.90 | < .001 | -4.62 | [-4.73, -4.52] |
| Weeks-Since-Birth (WSB) | 64.53 | < .001 | 0.03 | [0.03, 0.03] |
| Sex | -1.78 | .076 | -0.06 | [-0.13, 0.01] |
| Maximum Caregiver Education | -1.35 | .178 | -0.04 | [-0.11, 0.02] |
| Risk of Lead Exposure | 2.24 | .025 | 0.03 | [0.004, 0.05] |
| WSB × Risk of Lead Exposure | -0.32 | .751 | -0.0001 | [-0.0004, 0.0003] |

**Note**: The linear mixed-effects model incorporates testing the statistical significance of coefficients against a *t*-distribution. Sex was a categorical factor, effect coded with Male/Female as ‑1/+1. WSB, maximum caregiver education and risk of lead exposure were centered, continuous factors. The random-effects structure included separate random intercepts for participant ID number and for the participant’s study site. Tooth-lead levels were operationally defined relative to calcium (Ca) content in each tooth (^208^Pb:^43^Ca ratio) to control for overall mineral content. Analysis included 8,167 data points. The model accounted for 82.2% of the variance in the data (*R*^2^ = .822, adjusted *R*^2^ = .822).

**Table S11. Linear mixed-effects model output for the analysis of tooth-lead levels from +13 to +43 weeks since birth (WSB), with sex, household income, weeks-since-birth (WSB), risk of Pb exposure, and WSB × Risk of Pb Exposure as predictors.**

|  | *t*(8161) | *p* | *b* | 95% CI |
| --- | --- | --- | --- | --- |
| Intercept | -92.44 | < .001 | -4.63 | [-4.73, -4.53] |
| Weeks-Since-Birth (WSB) | 64.55 | < .001 | 0.03 | [0.03, 0.03] |
| Sex | -1.81 | .070 | -0.06 | [-0.13, 0.01] |
| Household Income | -1.43 | .152 | -0.02 | [-0.05, 0.01] |
| Risk of Lead Exposure | 2.28 | .022 | 0.03 | [0.004, 0.05] |
| WSB × Risk of Lead Exposure | -0.31 | .754 | -0.0001 | [-0.0004, 0.0003] |

**Note**: The linear mixed-effects model incorporates testing the statistical significance of coefficients against a *t*-distribution. Sex was a categorical factor, effect coded with Male/Female as ‑1/+1. WSB, household income, and risk of lead exposure were centered, continuous factors. The random-effects structure included separate random intercepts for participant ID number and for the participant’s study site. Tooth-lead levels were operationally defined relative to calcium (Ca) content in each tooth (^208^Pb:^43^Ca ratio) to control for overall mineral content. Analysis included 8,167 data points. The model accounted for 82.2% of the variance in the data (*R*^2^ = .822, adjusted *R*^2^ = .822).

**Table S92. Linear mixed-effects model output for the total-cognition performance (per the NIH Toolbox) as a function of tooth-lead levels from -12 to +13 weeks since birth (WSB), controlling for sex, age, risk of lead exposure, household income, and WSB.**

|  | *t*(11291) | *p* | *b* | 95% CI |
| --- | --- | --- | --- | --- |
| Intercept | 171.81 | < .001 | 87.25 | [86.26, 88.25] |
| Weeks-Since-Birth (WSB) | -0.03 | .976 | -0.0004 | [-0.02, 0.02] |
| Sex | 5.68 | < .001 | 0.41 | [0.27, 0.55] |
| Risk of Lead Exposure | 0.99 | .322 | 0.03 | [-0.03, 0.08] |
| Tooth-Lead Levels | -3.63 | < .001 | -0.46 | [-0.71, -0.21] |
| Household Income | 19.55 | < .001 | 0.71 | [0.64, 0.79] |
| Age | 43.31 | < .001 | 5.01 | [4.78, 5.24] |
| WSB × Tooth-Lead Levels | -0.64 | .524 | -0.02 | [-0.09, 0.05] |
| Tooth-Lead Levels × Household Income | 0.80 | .426 | 0.04 | [-0.06, 0.13] |
| Sex × Age | 2.46 | .014 | 0.28 | [0.06, 0.51] |

**Note**: The linear mixed-effects model incorporates testing the statistical significance of coefficients against a *t*-distribution. Sex was a categorical factor, effect coded with Male/Female as ‑1/+1. WSB, risk of lead exposure, tooth-lead levels, household income, and age were centered, continuous factors. Tooth-lead levels were log-transformed prior to centering. The random-effects structure included a random intercept for the participant’s study site, and WSB-varying tooth-lead levels by participant (WSB:Tooth-Lead-Levels interaction minus the intercept and lower-order terms). Tooth-lead levels were operationally defined relative to calcium (Ca) content in each tooth (^208^Pb:^43^Ca ratio) to control for overall mineral content. Analysis included 11,301 data points across 440 participants. Results were considered statistically significant at a *p*-value of .005. The model accounted for 28.3% of the variance in the data (*R*^2^ = .283, adjusted *R*^2^ = .282).

**Table S93. Linear mixed-effects model output for the crystallized-cognition performance (per the NIH Toolbox) as a function of tooth-lead levels from -12 to +13 weeks since birth (WSB), controlling for sex, age, risk of lead exposure, household income, and WSB.**

|  | *t*(11390) | *p* | *b* | 95% CI |
| --- | --- | --- | --- | --- |
| Intercept | 214.29 | < .001 | 87.04 | [86.25, 87.84] |
| Weeks-Since-Birth (WSB) | -1.02 | .306 | -0.01 | [-0.03, 0.01] |
| Sex | -0.75 | .452 | -0.04 | [-0.16, 0.07] |
| Risk of Lead Exposure | -0.60 | .546 | -0.01 | [-0.06, 0.03] |
| Tooth-Lead Levels | -4.85 | < .001 | -0.50 | [-0.70, -0.30] |
| Household Income | 19.56 | < .001 | 0.59 | [0.53, 0.65] |
| Age | 35.58 | < .001 | 3.37 | [3.18, 3.55] |
| WSB × Tooth-Lead Levels | -0.93 | .350 | -0.03 | [-0.08, 0.03] |
| Tooth-Lead Levels × Household Income | 3.94 | < .001 | 0.15 | [0.08, 0.23] |
| Sex × Age | -1.55 | .121 | -0.15 | [-0.33, 0.04] |

**Note**: The linear mixed-effects model incorporates testing the statistical significance of coefficients against a *t*-distribution. Sex was a categorical factor, effect coded with Male/Female as ‑1/+1. WSB, risk of lead exposure, tooth-lead levels, household income, and age were centered, continuous factors. Tooth-lead levels were log-transformed prior to centering. The random-effects structure included a random intercept for the participant’s study site, and WSB-varying tooth-lead levels by participant (WSB:Tooth-Lead-Levels interaction minus the intercept and lower-order terms). Tooth-lead levels were operationally defined relative to calcium (Ca) content in each tooth (^208^Pb:^43^Ca ratio) to control for overall mineral content. Analysis included 11,400 data points across 444 participants. Results were considered statistically significant at a *p*-value of .005. The model accounted for 25.3% of the variance in the data (*R*^2^ = .253, adjusted *R*^2^ = .252).

**Table S94. Linear mixed-effects model output for the fluid-cognition performance (per the NIH Toolbox) as a function of tooth-lead levels from -12 to +13 weeks since birth (WSB), controlling for sex, age, risk of lead exposure, household income, and WSB.**

|  | *t*(11291) | *p* | *b* | 95% CI |
| --- | --- | --- | --- | --- |
| Intercept | 176.05 | < .001 | 92.59 | [91.56, 93.63] |
| Weeks-Since-Birth (WSB) | 0.45 | .650 | 0.01 | [-0.02, 0.03] |
| Sex | 8.82 | < .001 | 0.76 | [0.59, 0.93] |
| Risk of Lead Exposure | 2.12 | .034 | 0.07 | [0.01, 0.13] |
| Tooth-Lead Levels | -1.57 | .116 | -0.24 | [-0.53, 0.06] |
| Household Income | 13.50 | < .001 | 0.59 | [0.50, 0.67] |
| Age | 35.43 | < .001 | 4.89 | [4.62, 5.16] |
| WSB × Tooth-Lead Levels | -0.47 | .640 | -0.02 | [-0.10, 0.06] |
| Tooth-Lead Levels × Household Income | -1.80 | .072 | -0.10 | [-0.21, 0.01] |
| Sex × Age | 5.22 | < .001 | 0.71 | [0.45, 0.98] |

**Note**: The linear mixed-effects model incorporates testing the statistical significance of coefficients against a *t*-distribution. Sex was a categorical factor, effect coded with Male/Female as ‑1/+1. WSB, risk of lead exposure, tooth-lead levels, household income, and age were centered, continuous factors. Tooth-lead levels were log-transformed prior to centering. The random-effects structure included a random intercept for the participant’s study site, and WSB-varying tooth-lead levels by participant (WSB:Tooth-Lead-Levels interaction minus the intercept and lower-order terms). Tooth-lead levels were operationally defined relative to calcium (Ca) content in each tooth (^208^Pb:^43^Ca ratio) to control for overall mineral content. Analysis included 11,301 data points across 440 participants. Results were considered statistically significant at a *p*-value of .005. The model accounted for 22.1% of the variance in the data (*R*^2^ = .221, adjusted *R*^2^ = .220).

**Table S95. Linear mixed-effects model output for the picture-vocabulary performance (per the NIH Toolbox) as a function of tooth-lead levels from -12 to +13 weeks since birth (WSB), controlling for sex, age, risk of lead exposure, household income, and WSB.**

|  | *t*(11390) | *p* | *b* | 95% CI |
| --- | --- | --- | --- | --- |
| Intercept | 171.71 | < .001 | 85.04 | [84.07, 86.01] |
| Weeks-Since-Birth (WSB) | -0.85 | .397 | -0.01 | [-0.03, 0.01] |
| Sex | 1.25 | .212 | 0.08 | [-0.05, 0.22] |
| Risk of Lead Exposure | -3.08 | .002 | -0.08 | [-0.13, -0.03] |
| Tooth-Lead Levels | -8.76 | < .001 | -1.04 | [-1.27, -0.81] |
| Household Income | 18.79 | < .001 | 0.65 | [0.58, 0.71] |
| Age | 33.71 | < .001 | 3.65 | [3.44, 3.87] |
| WSB × Tooth-Lead Levels | -0.07 | .948 | -0.002 | [-0.06, 0.06] |
| Tooth-Lead Levels × Household Income | 7.68 | < .001 | 0.34 | [0.26, 0.43] |
| Sex × Age | -4.95 | < .001 | -0.53 | [-0.74, -0.32] |

**Note**: The linear mixed-effects model incorporates testing the statistical significance of coefficients against a *t*-distribution. Sex was a categorical factor, effect coded with Male/Female as ‑1/+1. WSB, risk of lead exposure, tooth-lead levels, household income, and age were centered, continuous factors. Tooth-lead levels were log-transformed prior to centering. The random-effects structure included a random intercept for the participant’s study site, and WSB-varying tooth-lead levels by participant (WSB:Tooth-Lead-Levels interaction minus the intercept and lower-order terms). Tooth-lead levels were operationally defined relative to calcium (Ca) content in each tooth (^208^Pb:^43^Ca ratio) to control for overall mineral content. Analysis included 11,400 data points across 444 participants. Results were considered statistically significant at a *p*-value of .005. The model accounted for 25.3% of the variance in the data (*R*^2^ = .253, adjusted *R*^2^ = .253).

**Table S96. Linear mixed-effects model output for the oral-reading performance (per the NIH Toolbox) as a function of tooth-lead levels from -12 to +13 weeks since birth (WSB), controlling for sex, age, risk of lead exposure, household income, and WSB.**

|  | *t*(11390) | *p* | *b* | 95% CI |
| --- | --- | --- | --- | --- |
| Intercept | 251.25 | < .001 | 91.57 | [90.86, 92.29] |
| Weeks-Since-Birth (WSB) | -0.72 | .472 | -0.01 | [-0.03, 0.01] |
| Sex | -2.86 | .004 | -0.17 | [-0.28, -0.05] |
| Risk of Lead Exposure | 2.98 | .003 | 0.07 | [0.02, 0.11] |
| Tooth-Lead Levels | 0.52 | .600 | 0.05 | [-0.14, 0.25] |
| Household Income | 15.12 | < .001 | 0.45 | [0.39, 0.51] |
| Age | 28.13 | < .001 | 2.65 | [2.46, 2.83] |
| WSB × Tooth-Lead Levels | -1.44 | .150 | -0.05 | [-0.11, 0.02] |
| Tooth-Lead Levels × Household Income | -1.82 | .069 | -0.07 | [-0.15, 0.01] |
| Sex × Age | 2.65 | .008 | 0.25 | [0.06, 0.43] |

**Note**: The linear mixed-effects model incorporates testing the statistical significance of coefficients against a *t*-distribution. Sex was a categorical factor, effect coded with Male/Female as ‑1/+1. WSB, risk of lead exposure, tooth-lead levels, household income, and age were centered, continuous factors. Tooth-lead levels were log-transformed prior to centering. The random-effects structure included a random intercept for the participant’s study site, and WSB-varying tooth-lead levels by participant (WSB:Tooth-Lead-Levels interaction minus the intercept and lower-order terms). Tooth-lead levels were operationally defined relative to calcium (Ca) content in each tooth (^208^Pb:^43^Ca ratio) to control for overall mineral content. Analysis included 11,400 data points across 444 participants. Results were considered statistically significant at a *p*-value of .005. The model accounted for 20.5% of the variance in the data (*R*^2^ = .205, adjusted *R*^2^ = .204).

**Table S97. Linear mixed-effects model output for the total behavioral problems (per the Child Behavior Checklist) as a function of tooth-lead levels from -12 to +13 weeks since birth (WSB), controlling for sex, age, risk of lead exposure, household income, and WSB.**

|  | *t*(11492) | *p* | *b* | 95% CI |
| --- | --- | --- | --- | --- |
| Intercept | 59.22 | < .001 | 46.57 | [45.02, 48.11] |
| Weeks-Since-Birth (WSB) | 0.09 | .932 | 0.001 | [-0.03, 0.03] |
| Sex | -6.10 | < .001 | -0.60 | [-0.80, -0.41] |
| Risk of Lead Exposure | -1.87 | .062 | -0.07 | [-0.14, 0.003] |
| Tooth-Lead Levels | 3.03 | .002 | 0.52 | [0.18, 0.86] |
| Household Income | -6.12 | < .001 | -0.31 | [-0.40, -0.21] |
| Age | 0.47 | .640 | 0.07 | [-0.23, 0.38] |
| WSB × Tooth-Lead Levels | -0.67 | .505 | -0.03 | [-0.13, 0.06] |
| Tooth-Lead Levels × Household Income | 2.49 | .013 | 0.16 | [0.03, 0.29] |
| Sex × Age | -7.85 | < .001 | -1.22 | [-1.53, -0.92] |

**Note**: The linear mixed-effects model incorporates testing the statistical significance of coefficients against a *t*-distribution. Sex was a categorical factor, effect coded with Male/Female as ‑1/+1. WSB, risk of lead exposure, tooth-lead levels, household income, and age were centered, continuous factors. Tooth-lead levels were log-transformed prior to centering. The random-effects structure included a random intercept for the participant’s study site, and WSB-varying tooth-lead levels by participant (WSB:Tooth-Lead-Levels interaction minus the intercept and lower-order terms). Tooth-lead levels were operationally defined relative to calcium (Ca) content in each tooth (^208^Pb:^43^Ca ratio) to control for overall mineral content. Analysis included 11,502 data points across 448 participants. Results were considered statistically significant at a *p*-value of .005. The model accounted for 18.4% of the variance in the data (*R*^2^ = .184, adjusted *R*^2^ = .184).

**Table S98. Linear mixed-effects model output for the externalizing behavioral problems (per the Child Behavior Checklist) as a function of tooth-lead levels from -12 to +13 weeks since birth (WSB), controlling for sex, age, risk of lead exposure, household income, and WSB.**

|  | *t*(11492) | *p* | *b* | 95% CI |
| --- | --- | --- | --- | --- |
| Intercept | 66.32 | < .001 | 46.14 | [44.78, 47.51] |
| Weeks-Since-Birth (WSB) | -0.45 | .651 | -0.01 | [-0.04, 0.02] |
| Sex | -2.27 | .023 | -0.20 | [-0.38, -0.03] |
| Risk of Lead Exposure | 3.22 | .001 | 0.11 | [0.04, 0.18] |
| Tooth-Lead Levels | 2.76 | .006 | 0.43 | [0.13, 0.74] |
| Household Income | -5.88 | < .001 | -0.27 | [-0.36, -0.18] |
| Age | -1.75 | .081 | -0.25 | [-0.53, 0.03] |
| WSB × Tooth-Lead Levels | -0.64 | .520 | -0.03 | [-0.12, 0.06] |
| Tooth-Lead Levels × Household Income | 4.23 | < .001 | 0.25 | [0.13, 0.37] |
| Sex × Age | -6.32 | < .001 | -0.90 | [-1.18, -0.62] |

**Note**: The linear mixed-effects model incorporates testing the statistical significance of coefficients against a *t*-distribution. Sex was a categorical factor, effect coded with Male/Female as ‑1/+1. WSB, risk of lead exposure, tooth-lead levels, household income, and age were centered, continuous factors. Tooth-lead levels were log-transformed prior to centering. The random-effects structure included a random intercept for the participant’s study site, and WSB-varying tooth-lead levels by participant (WSB:Tooth-Lead-Levels interaction minus the intercept and lower-order terms). Tooth-lead levels were operationally defined relative to calcium (Ca) content in each tooth (^208^Pb:^43^Ca ratio) to control for overall mineral content. Analysis included 11,502 data points across 448 participants. Results were considered statistically significant at a *p*-value of .005. The model accounted for 17.1% of the variance in the data (*R*^2^ = .171, adjusted *R*^2^ = .170).

**Table S99. Linear mixed-effects model output for the internalizing behavioral problems (per the Child Behavior Checklist) as a function of tooth-lead levels from -12 to +13 weeks since birth (WSB), controlling for sex, age, risk of lead exposure, household income, and WSB.**

|  | *t*(11492) | *p* | *b* | 95% CI |
| --- | --- | --- | --- | --- |
| Intercept | 65.88 | < .001 | 49.22 | [47.76, 50.69] |
| Weeks-Since-Birth (WSB) | 0.86 | .388 | 0.01 | [-0.02, 0.04] |
| Sex | -8.80 | < .001 | -0.84 | [-1.02, -0.65] |
| Risk of Lead Exposure | -5.21 | < .001 | -0.19 | [-0.26, -0.12] |
| Tooth-Lead Levels | 2.01 | .044 | 0.33 | [0.01, 0.66] |
| Household Income | -5.21 | < .001 | -0.25 | [-0.35, -0.16] |
| Age | 7.22 | < .001 | 1.09 | [0.79, 1.39] |
| WSB × Tooth-Lead Levels | -0.29 | .772 | -0.01 | [-0.11, 0.08] |
| Tooth-Lead Levels × Household Income | 2.06 | .039 | 0.13 | [0.01, 0.25] |
| Sex × Age | -3.45 | .001 | -0.52 | [-0.81, -0.22] |

**Note**: The linear mixed-effects model incorporates testing the statistical significance of coefficients against a *t*-distribution. Sex was a categorical factor, effect coded with Male/Female as ‑1/+1. WSB, risk of lead exposure, tooth-lead levels, household income, and age were centered, continuous factors. Tooth-lead levels were log-transformed prior to centering. The random-effects structure included a random intercept for the participant’s study site, and WSB-varying tooth-lead levels by participant (WSB:Tooth-Lead-Levels interaction minus the intercept and lower-order terms). Tooth-lead levels were operationally defined relative to calcium (Ca) content in each tooth (^208^Pb:^43^Ca ratio) to control for overall mineral content. Analysis included 11,502 data points across 448 participants. Results were considered statistically significant at a *p*-value of .005. The model accounted for 18.9% of the variance in the data (*R*^2^ = .189, adjusted *R*^2^ = .189).

**Table S100. Linear mixed-effects model output for the picture-vocabulary performance (per the NIH Toolbox) as a function of tooth-lead levels from -12 to +0 weeks since birth (WSB) (i.e., prenatal exposure), controlling for sex, age, risk of lead exposure, household income, and WSB.**

|  | *t*(5649) | *p* | *b* | 95% CI |
| --- | --- | --- | --- | --- |
| Intercept | 160.99 | < .001 | 85.01 | [83.98, 86.05] |
| Weeks-Since-Birth (WSB) | 0.05 | .961 | 0.001 | [-0.04, 0.05] |
| Sex | -1.21 | .226 | -0.12 | [-0.30, 0.07] |
| Risk of Lead Exposure | -1.23 | .221 | -0.04 | [-0.11, 0.03] |
| Tooth-Lead Levels | -3.68 | < .001 | -0.82 | [-1.25, -0.38] |
| Household Income | 14.10 | < .001 | 0.69 | [0.60, 0.79] |
| Age | 25.44 | < .001 | 3.88 | [3.58, 4.18] |
| WSB × Tooth-Lead Levels | 1.75 | .080 | 0.16 | [-0.02, 0.34] |
| Tooth-Lead Levels × Household Income | 3.90 | < .001 | 0.32 | [0.16, 0.48] |
| Sex × Age | -2.35 | .019 | -0.36 | [-0.66, -0.06] |

**Note**: The linear mixed-effects model incorporates testing the statistical significance of coefficients against a *t*-distribution. Sex was a categorical factor, effect coded with Male/Female as ‑1/+1. WSB, risk of lead exposure, tooth-lead levels, household income, and age were centered, continuous factors. Tooth-lead levels were log-transformed prior to centering. The random-effects structure included a random intercept for the participant’s study site, and WSB-varying tooth-lead levels by participant (WSB:Tooth-Lead-Levels interaction minus the intercept and lower-order terms). Tooth-lead levels were operationally defined relative to calcium (Ca) content in each tooth (^208^Pb:^43^Ca ratio) to control for overall mineral content. Analysis included 5,659 data points across 444 participants. Results were considered statistically significant at a *p*-value of .005. The model accounted for 58.3% of the variance in the data (*R*^2^ = .583, adjusted *R*^2^ = .582).

**Table S101. Linear mixed-effects model output for the picture-vocabulary performance (per the NIH Toolbox) as a function of tooth-lead levels from +1 to +13 weeks since birth (WSB) (i.e., postnatal exposure), controlling for sex, age, risk of lead exposure, household income, and WSB.**

|  | *t*(5731) | *p* | *b* | 95% CI |
| --- | --- | --- | --- | --- |
| Intercept | 149.78 | < .001 | 84.95 | [83.84, 86.07] |
| Weeks-Since-Birth (WSB) | -0.38 | .701 | -0.01 | [-0.05, 0.03] |
| Sex | -1.96 | .050 | -0.18 | [-0.37, 0.0004] |
| Risk of Lead Exposure | -0.48 | .632 | -0.02 | [-0.09, 0.05] |
| Tooth-Lead Levels | -3.41 | .001 | -0.81 | [-1.28, -0.35] |
| Household Income | 12.66 | < .001 | 0.62 | [0.52, 0.71] |
| Age | 23.41 | < .001 | 3.58 | [3.28, 3.88] |
| WSB × Tooth-Lead Levels | -0.99 | .320 | -0.07 | [-0.20, 0.07] |
| Tooth-Lead Levels × Household Income | 3.19 | .001 | 0.27 | [0.10, 0.44] |
| Sex × Age | -2.91 | .004 | -0.44 | [-0.73, -0.14] |

**Note**: The linear mixed-effects model incorporates testing the statistical significance of coefficients against a *t*-distribution. Sex was a categorical factor, effect coded with Male/Female as ‑1/+1. WSB, risk of lead exposure, tooth-lead levels, household income, and age were centered, continuous factors. Tooth-lead levels were log-transformed prior to centering. The random-effects structure included a random intercept for the participant’s study site, and WSB-varying tooth-lead levels by participant (WSB:Tooth-Lead-Levels interaction minus the intercept and lower-order terms). Tooth-lead levels were operationally defined relative to calcium (Ca) content in each tooth (^208^Pb:^43^Ca ratio) to control for overall mineral content. Analysis included 5,741 data points across 444 participants. Results were considered statistically significant at a *p*-value of .005. The model accounted for 58.4% of the variance in the data (*R*^2^ = .584, adjusted *R*^2^ = .584).

**Table S102. Linear mixed-effects model output for the picture-vocabulary performance (per the NIH Toolbox) as a function of tooth-lead levels from ‑12 to +13 weeks since birth (WSB), having removed multivariate outliers (i.e., datapoints with larger Cook’s distance values), controlling for sex, age, risk of lead exposure, household income, and WSB.**

|  | *t*(10553) | *p* | *b* | 95% CI |
| --- | --- | --- | --- | --- |
| Intercept | 191.60 | < .001 | 84.75 | [83.89, 85.62] |
| Weeks-Since-Birth (WSB) | -0.73 | .468 | -0.01 | [-0.03, 0.01] |
| Sex | -0.85 | .396 | -0.05 | [-0.17, 0.07] |
| Risk of Lead Exposure | -0.88 | .379 | -0.02 | [-0.07, 0.03] |
| Tooth-Lead Levels | -9.92 | < .001 | -1.26 | [-1.50, -1.01] |
| Household Income | 16.70 | < .001 | 0.53 | [0.47, 0.59] |
| Age | 28.26 | < .001 | 2.82 | [2.62, 3.02] |
| WSB × Tooth-Lead Levels | -0.09 | .930 | -0.003 | [-0.07, 0.06] |
| Tooth-Lead Levels × Household Income | 5.98 | < .001 | 0.27 | [0.18, 0.35] |
| Sex × Age | -3.42 | .001 | -0.34 | [-0.53, -0.14] |

**Note**: The linear mixed-effects model incorporates testing the statistical significance of coefficients against a *t*-distribution. Sex was a categorical factor, effect coded with Male/Female as ‑1/+1. WSB, risk of lead exposure, tooth-lead levels, household income, and age were centered, continuous factors. Tooth-lead levels were log-transformed prior to centering. The random-effects structure included a random intercept for the participant’s study site, and WSB-varying tooth-lead levels by participant (WSB:Tooth-Lead-Levels interaction minus the intercept and lower-order terms). Tooth-lead levels were operationally defined relative to calcium (Ca) content in each tooth (^208^Pb:^43^Ca ratio) to control for overall mineral content. Analysis included 10,563 data points across 433 participants. Results were considered statistically significant at a *p*-value of .005. The model accounted for 23.6% of the variance in the data (*R*^2^ = .236, adjusted *R*^2^ = .236).

**Table S103. Linear mixed-effects model output for the externalizing behavioral problems (per the Child Behavior Checklist) as a function of tooth-lead levels from -12 to 0 weeks since birth (WSB) (i.e., prenatal exposure), controlling for sex, age, risk of lead exposure, household income, and WSB.**

|  | *t*(5699) | *p* | *b* | 95% CI |
| --- | --- | --- | --- | --- |
| Intercept | 57.45 | < .001 | 46.56 | [44.97, 48.14] |
| Weeks-Since-Birth (WSB) | -0.19 | .849 | -0.01 | [-0.07, 0.05] |
| Sex | -1.14 | .254 | -0.14 | [-0.39, 0.10] |
| Risk of Lead Exposure | 3.12 | .002 | 0.14 | [0.05, 0.24] |
| Tooth-Lead Levels | 0.72 | .471 | 0.21 | [-0.36, 0.79] |
| Household Income | -7.60 | < .001 | -0.50 | [-0.63, -0.37] |
| Age | 1.11 | .268 | 0.22 | [-0.17, 0.62] |
| WSB × Tooth-Lead Levels | -0.15 | .880 | -0.02 | [-0.28, 0.24] |
| Tooth-Lead Levels × Household Income | 1.99 | .046 | 0.22 | [0.003, 0.43] |
| Sex × Age | -3.29 | .001 | -0.66 | [-1.06, -0.27] |

**Note**: The linear mixed-effects model incorporates testing the statistical significance of coefficients against a *t*-distribution. Sex was a categorical factor, effect coded with Male/Female as ‑1/+1. WSB, risk of lead exposure, tooth-lead levels, household income, and age were centered, continuous factors. Tooth-lead levels were log-transformed prior to centering. The random-effects structure included a random intercept for the participant’s study site, and WSB-varying tooth-lead levels by participant (WSB:Tooth-Lead-Levels interaction minus the intercept and lower-order terms). Tooth-lead levels were operationally defined relative to calcium (Ca) content in each tooth (^208^Pb:^43^Ca ratio) to control for overall mineral content. Analysis included 5,709 data points across 448 participants. Results were considered statistically significant at a *p*-value of .005. The model accounted for 53.4% of the variance in the data (*R*^2^ = .534, adjusted *R*^2^ = .533).

**Table S104. Linear mixed-effects model output for the externalizing behavioral problems (per the Child Behavior Checklist) as a function of tooth-lead levels from +1 to +13 weeks since birth (WSB) (i.e., postnatal exposure), controlling for sex, age, risk of lead exposure, household income, and WSB.**

|  | *t*(5783) | *p* | *b* | 95% CI |
| --- | --- | --- | --- | --- |
| Intercept | 55.73 | < .001 | 46.54 | [44.91, 48.18] |
| Weeks-Since-Birth (WSB) | 0.27 | .789 | 0.01 | [-0.05, 0.07] |
| Sex | -1.70 | .090 | -0.22 | [-0.47, 0.03] |
| Risk of Lead Exposure | 1.19 | .233 | 0.06 | [-0.04, 0.15] |
| Tooth-Lead Levels | 1.40 | .162 | 0.45 | [-0.18, 1.09] |
| Household Income | -6.54 | < .001 | -0.43 | [-0.56, -0.30] |
| Age | 3.41 | .001 | 0.70 | [0.30, 1.11] |
| WSB × Tooth-Lead Levels | -1.09 | .277 | -0.10 | [-0.27, 0.08] |
| Tooth-Lead Levels × Household Income | 1.60 | .110 | 0.19 | [-0.04, 0.41] |
| Sex × Age | -4.92 | < .001 | -1.00 | [-1.40, -0.60] |

**Note**: The linear mixed-effects model incorporates testing the statistical significance of coefficients against a *t*-distribution. Sex was a categorical factor, effect coded with Male/Female as ‑1/+1. WSB, risk of lead exposure, tooth-lead levels, household income, and age were centered, continuous factors. Tooth-lead levels were log-transformed prior to centering. The random-effects structure included a random intercept for the participant’s study site, and WSB-varying tooth-lead levels by participant (WSB:Tooth-Lead-Levels interaction minus the intercept and lower-order terms). Tooth-lead levels were operationally defined relative to calcium (Ca) content in each tooth (^208^Pb:^43^Ca ratio) to control for overall mineral content. Analysis included 5,793 data points across 448 participants. Results were considered statistically significant at a *p*-value of .005. The model accounted for 50.7% of the variance in the data (*R*^2^ = .507, adjusted *R*^2^ = .507).

**Table S105. Linear mixed-effects model output for the externalizing behavioral problems (per the Child Behavior Checklist) as a function of tooth-lead levels from -12 to +13 weeks since birth (WSB), having removed multivariate outliers (i.e., datapoints with larger Cook’s distance values), controlling for sex, age, risk of lead exposure, household income, and WSB.**

|  | *t*(10741) | *p* | *b* | 95% CI |
| --- | --- | --- | --- | --- |
| Intercept | 66.81 | < .001 | 45.11 | [43.79, 46.44] |
| Weeks-Since-Birth (WSB) | -0.64 | .523 | -0.01 | [-0.03, 0.02] |
| Sex | 0.61 | .540 | 0.05 | [-0.11, 0.20] |
| Risk of Lead Exposure | 2.23 | .026 | 0.07 | [0.01, 0.12] |
| Tooth-Lead Levels | 1.73 | .084 | 0.26 | [-0.03, 0.56] |
| Household Income | -5.81 | < .001 | -0.23 | [-0.31, -0.16] |
| Age | 1.53 | .126 | 0.19 | [-0.05, 0.44] |
| WSB × Tooth-Lead Levels | 0.37 | .709 | 0.02 | [-0.08, 0.12] |
| Tooth-Lead Levels × Household Income | 2.69 | .007 | 0.15 | [0.04, 0.26] |
| Sex × Age | -5.55 | < .001 | -0.70 | [-0.95, -0.45] |

**Note**: The linear mixed-effects model incorporates testing the statistical significance of coefficients against a *t*-distribution. Sex was a categorical factor, effect coded with Male/Female as ‑1/+1. WSB, risk of lead exposure, tooth-lead levels, household income, and age were centered, continuous factors. Tooth-lead levels were log-transformed prior to centering. The random-effects structure included a random intercept for the participant’s study site, and WSB-varying tooth-lead levels by participant (WSB:Tooth-Lead-Levels interaction minus the intercept and lower-order terms). Tooth-lead levels were operationally defined relative to calcium (Ca) content in each tooth (^208^Pb:^43^Ca ratio) to control for overall mineral content. Analysis included 10,751 data points across 432 participants. Results were considered statistically significant at a *p*-value of .005. The model accounted for 20.3% of the variance in the data (*R*^2^ = .203, adjusted *R*^2^ = .202).

**Table S106. Adolescent Brain Cognitive Development (ABCD) Study Data Dictionary.**

| Data Release Variable Name | Data Description |
| --- | --- |
| **Shed Deciduous (“Baby”) Teeth** |  |
| age_wk | Weeks-Since-Birth (WSB) |
| tooth | Tooth Type |
| pb208 | Tooth-Lead Levels (Relative to Calcium Levels) |
| *baby_teeth_count (ph_p_teeth) | Total Number of Teeth Donated |
| **ABCD Standard Variables & Demographics** |  |
| ab_g_dyn__visit_age | Youth's age at the start of the event |
| ab_g_dyn__design_site | ABCD Study assessment site |
| ab_g_stc__cohort_ethnrace__leg | Ethno-racial identity (Legacy ABCD variable reporting 5 levels, used for recruitment targets) |
| ab_g_stc__cohort_sex | Youth participant's sex |
| ab_g_dyn__cohort_income__hhold__3lvl | Household income - 3 levels |
| ab_p_demo__income__hhold_001 | Total combined family income for the past 12 months |
| ab_g_dyn__cohort_edu__cgs | Highest education across caregivers |
| **Structural Magnetic Resonance Imaging (Cortical Thickness)** |  |
| mr_y_smri__thk__dsk__bstmps__lh_mean | Average cortical thickness in Desikan ROI: Banks of superior temporal sulcus (Left hemisphere) |
| mr_y_smri__thk__dsk__cac__lh_mean | Average cortical thickness in Desikan ROI: Caudal anterior cingulate (Left hemisphere) |
| mr_y_smri__thk__dsk__cmfrt__lh_mean | Average cortical thickness in Desikan ROI: Caudal middle frontal (Left hemisphere) |
| mr_y_smri__thk__dsk__cn__lh_mean | Average cortical thickness in Desikan ROI: Cuneus (Left hemisphere) |
| mr_y_smri__thk__dsk__er__lh_mean | Average cortical thickness in Desikan ROI: Entorhinal (Left hemisphere) |
| mr_y_smri__thk__dsk__ff__lh_mean | Average cortical thickness in Desikan ROI: Fusiform (Left hemisphere) |
| mr_y_smri__thk__dsk__ic__lh_mean | Average cortical thickness in Desikan ROI: Isthmus cingulate (Left hemisphere) |
| mr_y_smri__thk__dsk__ins__lh_mean | Average cortical thickness in Desikan ROI: Insula (Left hemisphere) |
| mr_y_smri__thk__dsk__iprt__lh_mean | Average cortical thickness in Desikan ROI: Inferior parietal (Left hemisphere) |
| mr_y_smri__thk__dsk__itmp__lh_mean | Average cortical thickness in Desikan ROI: Inferior temporal (Left hemisphere) |
| mr_y_smri__thk__dsk__lg__lh_mean | Average cortical thickness in Desikan ROI: Lingual (Left hemisphere) |
| mr_y_smri__thk__dsk__lobfrt__lh_mean | Average cortical thickness in Desikan ROI: Lateral orbitofrontal (Left hemisphere) |
| mr_y_smri__thk__dsk__locc__lh_mean | Average cortical thickness in Desikan ROI: Lateral occipital (Left hemisphere) |
| mr_y_smri__thk__dsk__mobfrt__lh_mean | Average cortical thickness in Desikan ROI: Medial orbitofrontal (Left hemisphere) |
| mr_y_smri__thk__dsk__mtmp__lh_mean | Average cortical thickness in Desikan ROI: Middle temporal (Left hemisphere) |
| mr_y_smri__thk__dsk__pactr__lh_mean | Average cortical thickness in Desikan ROI: Paracentral (Left hemisphere) |
| mr_y_smri__thk__dsk__pcc__lh_mean | Average cortical thickness in Desikan ROI: Pericalcarine (Left hemisphere) |
| mr_y_smri__thk__dsk__pcg__lh_mean | Average cortical thickness in Desikan ROI: Posterior cingulate (Left hemisphere) |
| mr_y_smri__thk__dsk__pfrt__lh_mean | Average cortical thickness in Desikan ROI: Frontal pole (Left hemisphere) |
| mr_y_smri__thk__dsk__ph__lh_mean | Average cortical thickness in Desikan ROI: Parahippocampal (Left hemisphere) |
| mr_y_smri__thk__dsk__pob__lh_mean | Average cortical thickness in Desikan ROI: Pars orbitalis (Left hemisphere) |
| mr_y_smri__thk__dsk__poctr__lh_mean | Average cortical thickness in Desikan ROI: Postcentral (Left hemisphere) |
| mr_y_smri__thk__dsk__pop__lh_mean | Average cortical thickness in Desikan ROI: Pars opercularis (Left hemisphere) |
| mr_y_smri__thk__dsk__prcn__lh_mean | Average cortical thickness in Desikan ROI: Precuneus (Left hemisphere) |
| mr_y_smri__thk__dsk__prctr__lh_mean | Average cortical thickness in Desikan ROI: Precentral (Left hemisphere) |
| mr_y_smri__thk__dsk__ptg__lh_mean | Average cortical thickness in Desikan ROI: Pars triangularis (Left hemisphere) |
| mr_y_smri__thk__dsk__ptmp__lh_mean | Average cortical thickness in Desikan ROI: Temporal pole (Left hemisphere) |
| mr_y_smri__thk__dsk__rac__lh_mean | Average cortical thickness in Desikan ROI: Rostral anterior cingulate (Left hemisphere) |
| mr_y_smri__thk__dsk__rmfrt__lh_mean | Average cortical thickness in Desikan ROI: Rostral middle frontal (Left hemisphere) |
| mr_y_smri__thk__dsk__sfrt__lh_mean | Average cortical thickness in Desikan ROI: Superior frontal (Left hemisphere) |
| mr_y_smri__thk__dsk__sm__lh_mean | Average cortical thickness in Desikan ROI: Supramarginal (Left hemisphere) |
| mr_y_smri__thk__dsk__sprt__lh_mean | Average cortical thickness in Desikan ROI: Superior parietal (Left hemisphere) |
| mr_y_smri__thk__dsk__stmp__lh_mean | Average cortical thickness in Desikan ROI: Superior temporal (Left hemisphere) |
| mr_y_smri__thk__dsk__ttmp__lh_mean | Average cortical thickness in Desikan ROI: Transverse temporal (Left hemisphere) |
| mr_y_smri__thk__dsk__bstmps__rh_mean | Average cortical thickness in Desikan ROI: Banks of superior temporal sulcus (Right hemisphere) |
| mr_y_smri__thk__dsk__cac__rh_mean | Average cortical thickness in Desikan ROI: Caudal anterior cingulate (Right hemisphere) |
| mr_y_smri__thk__dsk__cmfrt__rh_mean | Average cortical thickness in Desikan ROI: Caudal middle frontal (Right hemisphere) |
| mr_y_smri__thk__dsk__cn__rh_mean | Average cortical thickness in Desikan ROI: Cuneus (Right hemisphere) |
| mr_y_smri__thk__dsk__er__rh_mean | Average cortical thickness in Desikan ROI: Entorhinal (Right hemisphere) |
| mr_y_smri__thk__dsk__ff__rh_mean | Average cortical thickness in Desikan ROI: Fusiform (Right hemisphere) |
| mr_y_smri__thk__dsk__ic__rh_mean | Average cortical thickness in Desikan ROI: Isthmus cingulate (Right hemisphere) |
| mr_y_smri__thk__dsk__ins__rh_mean | Average cortical thickness in Desikan ROI: Insula (Right hemisphere) |
| mr_y_smri__thk__dsk__iprt__rh_mean | Average cortical thickness in Desikan ROI: Inferior parietal (Right hemisphere) |
| mr_y_smri__thk__dsk__itmp__rh_mean | Average cortical thickness in Desikan ROI: Inferior temporal (Right hemisphere) |
| mr_y_smri__thk__dsk__lg__rh_mean | Average cortical thickness in Desikan ROI: Lingual (Right hemisphere) |
| mr_y_smri__thk__dsk__lobfrt__rh_mean | Average cortical thickness in Desikan ROI: Lateral orbitofrontal (Right hemisphere) |
| mr_y_smri__thk__dsk__locc__rh_mean | Average cortical thickness in Desikan ROI: Lateral occipital (Right hemisphere) |
| mr_y_smri__thk__dsk__mobfrt__rh_mean | Average cortical thickness in Desikan ROI: Medial orbitofrontal (Right hemisphere) |
| mr_y_smri__thk__dsk__mtmp__rh_mean | Average cortical thickness in Desikan ROI: Middle temporal (Right hemisphere) |
| mr_y_smri__thk__dsk__pactr__rh_mean | Average cortical thickness in Desikan ROI: Paracentral (Right hemisphere) |
| mr_y_smri__thk__dsk__pcc__rh_mean | Average cortical thickness in Desikan ROI: Pericalcarine (Right hemisphere) |
| mr_y_smri__thk__dsk__pcg__rh_mean | Average cortical thickness in Desikan ROI: Posterior cingulate (Right hemisphere) |
| mr_y_smri__thk__dsk__pfrt__rh_mean | Average cortical thickness in Desikan ROI: Frontal pole (Right hemisphere) |
| mr_y_smri__thk__dsk__ph__rh_mean | Average cortical thickness in Desikan ROI: Parahippocampal (Right hemisphere) |
| mr_y_smri__thk__dsk__pob__rh_mean | Average cortical thickness in Desikan ROI: Pars orbitalis (Right hemisphere) |
| mr_y_smri__thk__dsk__poctr__rh_mean | Average cortical thickness in Desikan ROI: Postcentral (Right hemisphere) |
| mr_y_smri__thk__dsk__pop__rh_mean | Average cortical thickness in Desikan ROI: Pars opercularis (Right hemisphere) |
| mr_y_smri__thk__dsk__prcn__rh_mean | Average cortical thickness in Desikan ROI: Precuneus (Right hemisphere) |
| mr_y_smri__thk__dsk__prctr__rh_mean | Average cortical thickness in Desikan ROI: Precentral (Right hemisphere) |
| mr_y_smri__thk__dsk__ptg__rh_mean | Average cortical thickness in Desikan ROI: Pars triangularis (Right hemisphere) |
| mr_y_smri__thk__dsk__ptmp__rh_mean | Average cortical thickness in Desikan ROI: Temporal pole (Right hemisphere) |
| mr_y_smri__thk__dsk__rac__rh_mean | Average cortical thickness in Desikan ROI: Rostral anterior cingulate (Right hemisphere) |
| mr_y_smri__thk__dsk__rmfrt__rh_mean | Average cortical thickness in Desikan ROI: Rostral middle frontal (Right hemisphere) |
| mr_y_smri__thk__dsk__sfrt__rh_mean | Average cortical thickness in Desikan ROI: Superior frontal (Right hemisphere) |
| mr_y_smri__thk__dsk__sm__rh_mean | Average cortical thickness in Desikan ROI: Supramarginal (Right hemisphere) |
| mr_y_smri__thk__dsk__sprt__rh_mean | Average cortical thickness in Desikan ROI: Superior parietal (Right hemisphere) |
| mr_y_smri__thk__dsk__stmp__rh_mean | Average cortical thickness in Desikan ROI: Superior temporal (Right hemisphere) |
| mr_y_smri__thk__dsk__ttmp__rh_mean | Average cortical thickness in Desikan ROI: Transverse temporal (Right hemisphere) |
| **Structural Magnetic Resonance Imaging (Cortical Surface Area)** |  |
| mr_y_smri__area__dsk__bstmps__lh_sum | Total surface area of Desikan ROI: Banks of superior temporal sulcus (Left hemisphere) |
| mr_y_smri__area__dsk__cac__lh_sum | Total surface area of Desikan ROI: Caudal anterior cingulate (Left hemisphere) |
| mr_y_smri__area__dsk__cmfrt__lh_sum | Total surface area of Desikan ROI: Caudal middle frontal (Left hemisphere) |
| mr_y_smri__area__dsk__cn__lh_sum | Total surface area of Desikan ROI: Cuneus (Left hemisphere) |
| mr_y_smri__area__dsk__er__lh_sum | Total surface area of Desikan ROI: Entorhinal (Left hemisphere) |
| mr_y_smri__area__dsk__ff__lh_sum | Total surface area of Desikan ROI: Fusiform (Left hemisphere) |
| mr_y_smri__area__dsk__ic__lh_sum | Total surface area of Desikan ROI: Isthmus cingulate (Left hemisphere) |
| mr_y_smri__area__dsk__ins__lh_sum | Total surface area of Desikan ROI: Insula (Left hemisphere) |
| mr_y_smri__area__dsk__iprt__lh_sum | Total surface area of Desikan ROI: Inferior parietal (Left hemisphere) |
| mr_y_smri__area__dsk__itmp__lh_sum | Total surface area of Desikan ROI: Inferior temporal (Left hemisphere) |
| mr_y_smri__area__dsk__lg__lh_sum | Total surface area of Desikan ROI: Lingual (Left hemisphere) |
| mr_y_smri__area__dsk__lobfrt__lh_sum | Total surface area of Desikan ROI: Lateral orbitofrontal (Left hemisphere) |
| mr_y_smri__area__dsk__locc__lh_sum | Total surface area of Desikan ROI: Lateral occipital (Left hemisphere) |
| mr_y_smri__area__dsk__mobfrt__lh_sum | Total surface area of Desikan ROI: Medial orbitofrontal (Left hemisphere) |
| mr_y_smri__area__dsk__mtmp__lh_sum | Total surface area of Desikan ROI: Middle temporal (Left hemisphere) |
| mr_y_smri__area__dsk__pactr__lh_sum | Total surface area of Desikan ROI: Paracentral (Left hemisphere) |
| mr_y_smri__area__dsk__pcc__lh_sum | Total surface area of Desikan ROI: Pericalcarine (Left hemisphere) |
| mr_y_smri__area__dsk__pcg__lh_sum | Total surface area of Desikan ROI: Posterior cingulate (Left hemisphere) |
| mr_y_smri__area__dsk__pfrt__lh_sum | Total surface area of Desikan ROI: Frontal pole (Left hemisphere) |
| mr_y_smri__area__dsk__ph__lh_sum | Total surface area of Desikan ROI: Parahippocampal (Left hemisphere) |
| mr_y_smri__area__dsk__pob__lh_sum | Total surface area of Desikan ROI: Pars orbitalis (Left hemisphere) |
| mr_y_smri__area__dsk__poctr__lh_sum | Total surface area of Desikan ROI: Postcentral (Left hemisphere) |
| mr_y_smri__area__dsk__pop__lh_sum | Total surface area of Desikan ROI: Pars opercularis (Left hemisphere) |
| mr_y_smri__area__dsk__prcn__lh_sum | Total surface area of Desikan ROI: Precuneus (Left hemisphere) |
| mr_y_smri__area__dsk__prctr__lh_sum | Total surface area of Desikan ROI: Precentral (Left hemisphere) |
| mr_y_smri__area__dsk__ptg__lh_sum | Total surface area of Desikan ROI: Pars triangularis (Left hemisphere) |
| mr_y_smri__area__dsk__ptmp__lh_sum | Total surface area of Desikan ROI: Temporal pole (Left hemisphere) |
| mr_y_smri__area__dsk__rac__lh_sum | Total surface area of Desikan ROI: Rostral anterior cingulate (Left hemisphere) |
| mr_y_smri__area__dsk__rmfrt__lh_sum | Total surface area of Desikan ROI: Rostral middle frontal (Left hemisphere) |
| mr_y_smri__area__dsk__sfrt__lh_sum | Total surface area of Desikan ROI: Superior frontal (Left hemisphere) |
| mr_y_smri__area__dsk__sm__lh_sum | Total surface area of Desikan ROI: Supramarginal (Left hemisphere) |
| mr_y_smri__area__dsk__sprt__lh_sum | Total surface area of Desikan ROI: Superior parietal (Left hemisphere) |
| mr_y_smri__area__dsk__stmp__lh_sum | Total surface area of Desikan ROI: Superior temporal (Left hemisphere) |
| mr_y_smri__area__dsk__ttmp__lh_sum | Total surface area of Desikan ROI: Transverse temporal (Left hemisphere) |
| mr_y_smri__area__dsk__bstmps__rh_sum | Total surface area of Desikan ROI: Banks of superior temporal sulcus (Right hemisphere) |
| mr_y_smri__area__dsk__cac__rh_sum | Total surface area of Desikan ROI: Caudal anterior cingulate (Right hemisphere) |
| mr_y_smri__area__dsk__cmfrt__rh_sum | Total surface area of Desikan ROI: Caudal middle frontal (Right hemisphere) |
| mr_y_smri__area__dsk__cn__rh_sum | Total surface area of Desikan ROI: Cuneus (Right hemisphere) |
| mr_y_smri__area__dsk__er__rh_sum | Total surface area of Desikan ROI: Entorhinal (Right hemisphere) |
| mr_y_smri__area__dsk__ff__rh_sum | Total surface area of Desikan ROI: Fusiform (Right hemisphere) |
| mr_y_smri__area__dsk__ic__rh_sum | Total surface area of Desikan ROI: Isthmus cingulate (Right hemisphere) |
| mr_y_smri__area__dsk__ins__rh_sum | Total surface area of Desikan ROI: Insula (Right hemisphere) |
| mr_y_smri__area__dsk__iprt__rh_sum | Total surface area of Desikan ROI: Inferior parietal (Right hemisphere) |
| mr_y_smri__area__dsk__itmp__rh_sum | Total surface area of Desikan ROI: Inferior temporal (Right hemisphere) |
| mr_y_smri__area__dsk__lg__rh_sum | Total surface area of Desikan ROI: Lingual (Right hemisphere) |
| mr_y_smri__area__dsk__lobfrt__rh_sum | Total surface area of Desikan ROI: Lateral orbitofrontal (Right hemisphere) |
| mr_y_smri__area__dsk__locc__rh_sum | Total surface area of Desikan ROI: Lateral occipital (Right hemisphere) |
| mr_y_smri__area__dsk__mobfrt__rh_sum | Total surface area of Desikan ROI: Medial orbitofrontal (Right hemisphere) |
| mr_y_smri__area__dsk__mtmp__rh_sum | Total surface area of Desikan ROI: Middle temporal (Right hemisphere) |
| mr_y_smri__area__dsk__pactr__rh_sum | Total surface area of Desikan ROI: Paracentral (Right hemisphere) |
| mr_y_smri__area__dsk__pcc__rh_sum | Total surface area of Desikan ROI: Pericalcarine (Right hemisphere) |
| mr_y_smri__area__dsk__pcg__rh_sum | Total surface area of Desikan ROI: Posterior cingulate (Right hemisphere) |
| mr_y_smri__area__dsk__pfrt__rh_sum | Total surface area of Desikan ROI: Frontal pole (Right hemisphere) |
| mr_y_smri__area__dsk__ph__rh_sum | Total surface area of Desikan ROI: Parahippocampal (Right hemisphere) |
| mr_y_smri__area__dsk__pob__rh_sum | Total surface area of Desikan ROI: Pars orbitalis (Right hemisphere) |
| mr_y_smri__area__dsk__poctr__rh_sum | Total surface area of Desikan ROI: Postcentral (Right hemisphere) |
| mr_y_smri__area__dsk__pop__rh_sum | Total surface area of Desikan ROI: Pars opercularis (Right hemisphere) |
| mr_y_smri__area__dsk__prcn__rh_sum | Total surface area of Desikan ROI: Precuneus (Right hemisphere) |
| mr_y_smri__area__dsk__prctr__rh_sum | Total surface area of Desikan ROI: Precentral (Right hemisphere) |
| mr_y_smri__area__dsk__ptg__rh_sum | Total surface area of Desikan ROI: Pars triangularis (Right hemisphere) |
| mr_y_smri__area__dsk__ptmp__rh_sum | Total surface area of Desikan ROI: Temporal pole (Right hemisphere) |
| mr_y_smri__area__dsk__rac__rh_sum | Total surface area of Desikan ROI: Rostral anterior cingulate (Right hemisphere) |
| mr_y_smri__area__dsk__rmfrt__rh_sum | Total surface area of Desikan ROI: Rostral middle frontal (Right hemisphere) |
| mr_y_smri__area__dsk__sfrt__rh_sum | Total surface area of Desikan ROI: Superior frontal (Right hemisphere) |
| mr_y_smri__area__dsk__sm__rh_sum | Total surface area of Desikan ROI: Supramarginal (Right hemisphere) |
| mr_y_smri__area__dsk__sprt__rh_sum | Total surface area of Desikan ROI: Superior parietal (Right hemisphere) |
| mr_y_smri__area__dsk__stmp__rh_sum | Total surface area of Desikan ROI: Superior temporal (Right hemisphere) |
| mr_y_smri__area__dsk__ttmp__rh_sum | Total surface area of Desikan ROI: Transverse temporal (Right hemisphere) |
| **Structural Magnetic Resonance Imaging (Covariates, Scanner, and Quality Control)** |  |
| mr_y_smri__area__dsk_sum | Total surface area of Desikan ROI: All |
| mr_y_smri__vol__aseg__whb_sum | Whole-brain volume |
| mr_y_adm__info__dev_serial | Imaging device hashed serial number |
| mr_y_qc__incl__smri__t1_indicator | Structural MRI - T1 weighted: Data recommended for inclusion |
| **Risk of Lead Exposure** |  |
| le_l_leadrisk__addr1_idx | Primary address: Census-tract-level estimated lead risk (1-10 scale) |
| le_l_leadrisk__addr1__pov125_prcnt | Primary address: Census-tract-level percentage of individuals below -125 percent of poverty level for primary address |
| le_l_leadrisk__addr1__homerisk_prcnt | Primary address: Census-tract-level estimated percentage of homes at risk for lead exposure given lead-based paint |
| **Cognition and Behavior** |  |
| nc_y_nihtb__comp__tot__uncor_score | Youth’s cognition total composite - Uncorrected standard score (NIH Toolbox) |
| nc_y_nihtb__comp__cryst__uncor_score | Youth’s crystallized composite - Uncorrected standard score (NIH Toolbox) |
| nc_y_nihtb__comp__fluid__uncor_score | Youth’s fluid composite - Uncorrected standard score (NIH Toolbox) |
| nc_y_nihtb__picvcb__uncor_score | Youth’s Picture Vocabulary Task - Uncorrected Standard Score (NIH Toolbox) |
| nc_y_nihtb__readr__uncor_score | Youth’s Oral Reading Recognition Task - Uncorrected Standard Score (NIH Toolbox) |
| mh_p_cbcl_tscore | Child Behavior Checklist (per caregiver reports about youth participant): T-score |
| mh_p_cbcl__synd__ext_tscore | Child Behavior Checklist (per caregiver reports about youth participant): Externalizing T-score |
| mh_p_cbcl__synd__int_tscore | Child Behavior Checklist (per caregiver reports about youth participant): Internalizing T-score |

**Note**: *Except for the total teeth donated, all variable names refer to variables in the May 2026 ABCD 7.0 Data Release (doi: 10.82525/8f3w-5260; https://www.nbdc-datahub.org/abcd-release-7-0). The total-teeth-donated data were extracted from the November 2023 ABCD 5.1 data release (10.15154/z563-zd24) from the designated variable name and table.
